# Recognition and inhibition of SARS-CoV-2 by humoral innate immunity pattern recognition molecules

**DOI:** 10.1101/2021.06.07.21258350

**Authors:** Matteo Stravalaci, Isabel Pagani, Elvezia Maria Paraboschi, Mattia Pedotti, Andrea Doni, Francesco Scavello, Sarah N. Mapelli, Marina Sironi, Luca Varani, Milos Matkovic, Andrea Cavalli, Daniela Cesana, Pierangela Gallina, Nicoletta Pedemonte, Valeria Capurro, Nicola Clementi, Nicasio Mancini, Pietro Invernizzi, Rino Rappuoli, Stefano Duga, Barbara Bottazzi, Mariagrazia Uguccioni, Rosanna Asselta, Elisa Vicenzi, Alberto Mantovani, Cecilia Garlanda

**Affiliations:** IRCCS Humanitas Research Hospital, via Manzoni 56, 20089 Rozzano (Milan), Italy; Department of Biomedical Sciences, Humanitas University, Via Rita Levi Montalcini 4, 20090 Pieve Emanuele (Milan), Italy; Viral Pathogenesis and Biosafety Unit, IRCCS San Raffaele Scientific Institute, Milan, Italy; Institute for Research in Biomedicine, Università della Svizzera italiana (USI), Bellinzona, Switzerland; Swiss Institute of Bioinformatics, Lausanne, Switzerland; San Raffaele Telethon Institute for Gene Therapy (SR-Tiget); IRCCS, San Raffaele Scientific Institute, Milan, Italy; UOC Genetica Medica, IRCCS Istituto Giannina Gaslini, Via Gaslini 5, 16147 Genova; Laboratory of Microbiology and Virology, IRCCS Scientific Institute and Vita-Salute San Raffaele University, Milan Italy; Division of Gastroenterology, Center for Autoimmune Liver Diseases, Department of Medicine and Surgery, University of Milano-Bicocca, Monza, Italy; European Reference Network on Hepatological Diseases (ERN RARE-LIVER), San Gerardo Hospital, Monza, Italy; Monoclonal Antibody Discovery Lab, Fondazione Toscana Life Sciences, Siena, Italy; Faculty of Medicine, Imperial College London, London, UK; The William Harvey Research Institute, Queen Mary University of London, Charterhouse Square, London EC1M 6BQ.

## Abstract

The humoral arm of innate immunity includes diverse molecules with antibody-like functions, some of which serve as disease severity biomarkers in COVID-19. The present study was designed to conduct a systematic investigation of the interaction of humoral fluid phase pattern recognition molecules (PRM) with SARS-CoV-2. Out of 10 PRM tested, the long pentraxin PTX3 and Mannose Binding Lectin (MBL) bound the viral Nucleoprotein and Spike, respectively. MBL bound trimeric Spike, including that of variants of concern, in a glycan-dependent way and inhibited SARS-CoV-2 in three *in vitro* models. Moreover, upon binding to Spike, MBL activated the lectin pathway of complement activation. Genetic polymorphisms at the MBL locus were associated with disease severity. These results suggest that selected humoral fluid phase PRM can play an important role in resistance to, and pathogenesis of, COVID-19, a finding with translational implications.

## Introduction

SARS-CoV-2 is a highly pathogenic coronavirus and the causative agent of the current Coronavirus disease-2019 (COVID-19) pandemic (Hartenian et al., 2020; Wu et al., 2020; Zhu et al., 2020). Innate immunity is credited to play a fundamental role in this condition and may eradicate the infection in its early phases, before adaptive immune responses take place. In severe forms of the disease, uncontrolled activation of innate and adaptive immunity results in hyperinflammatory responses, which affect the lung and blood vessels, contributing to ARDS, shock and multiorgan failure (Wang et al., 2020).

Innate immunity includes a cellular and a humoral arm (Bottazzi et al., 2010). The humoral arm consists in soluble PRMs belonging to different families, which include collectins [e.g. mannose binding lectin (MBL)], ficolins, pentraxins [C-reactive protein (CRP), Serum amyloid P component (SAP), pentraxin 3 (PTX3)], and C1q (Bottazzi et al., 2010; Garlanda et al., 2018; Holmskov et al., 2003). Humoral PRMs represent functional ancestors of antibodies (ante-antibodies), as they recognize microbial components and eliminate pathogens with common mechanisms that include agglutination, neutralization, activation of the complement cascade and opsonization facilitating phagocytosis (Bottazzi et al., 2010). Investigations of the role of humoral innate immunity in viral sensing have shown that collectins bind to envelope glycoproteins on enveloped viruses, including influenza virus, human immunodeficiency virus, hepatitis C virus (HCV) and herpes simplex virus, as well as to the nonenveloped rotavirus (Holmskov et al., 2003). Interaction may result in opsonization, agglutination, inhibition of viral fusion and entry, or complement activation, generally leading to inhibition of infection (Holmskov et al., 2003). Among pentraxins, the long pentraxin PTX3 has been show to interact with H3N2-subtype influenza virus type A by interacting with viral envelope hemagglutinin and neuraminidase glycoproteins through a sialic acid residue on its glycosidic moiety (Reading et al., 2008), with cytomegalovirus (CMV) (Bozza et al., 2006), and with the coronavirus murine hepatitis virus strain 1 (MHV-1) (Han et al., 2012), preventing viral infection.

Several lines of evidence, including genetic associations, indicate that cellular innate immunity, and related cytokines and chemokines, play a key role in SARS-CoV-2 recognition, antiviral resistance and, at later stages, severe disease (Merad and Martin, 2020; Pairo- Castineira et al., 2021; Severe Covid et al., 2020; Zhang et al., 2020). In contrast, little information is available concerning the role of the humoral arm of innate immunity in COVID- 19 resistance and pathogenesis, in spite of the clinical prognostic significance of CRP and PTX3 (Brunetta et al., 2021; Fajgenbaum and June, 2020).

The present study was designed to conduct a systematic investigation of the interaction of the humoral PRM with SARS-CoV-2. We found that PTX3 and MBL bound the SARS-CoV-2 Nucleoprotein and Spike, respectively. MBL recognized variants of concern (VoC), had antiviral activity and activated the Complement lectin pathway. Genetic polymorphisms at the MBL locus were associated with disease severity. Thus, selected fluid phase PRM (ante- antibodies) play an important role in resistance to, and pathogenesis of, COVID-19, a finding with translational implications.

## Results

### Interaction of humoral pattern recognition molecules with SARS-CoV-2 proteins

To study the role of humoral PRMs in recognizing SARS-CoV-2, we first investigated the interaction between humoral innate immunity molecules and SARS-CoV-2 proteins using a solid phase binding assay. We first analysed pentraxins and, as shown in Figure 1A and 1B, we did not observe specific binding of CRP and SAP to any of the SARS-CoV-2 proteins tested (S1, S2, S protein active trimer, Nucleocapsid, Envelope protein). In contrast, PTX3 bound specifically and in a dose-dependent manner to the Nucleocapsid protein, one of the most abundant proteins of SARS-CoV-2 (Zeng et al., 2020) (Figure 1C). We validated this result by confirming the binding of PTX3 with SARS-CoV-2 Nucleocapsid protein produced obtained from different sources. PTX3 is a multimeric glycoprotein arranged in an octameric structure. Each protomer is constituted of a flexible N-terminal region and a C-terminal domain with homology to the short pentraxin family (Bottazzi et al., 2010). To define which portion of the molecule was involved in the interaction, we compared the binding of full length PTX3 and its N-terminal or C-terminal domains to SARS-CoV-2 Nucleocapsid protein. Results indicate that PTX3 interacts with SARS-CoV-2 Nucleocapsid protein mainly through its N-terminal domain, although with lower affinity compared to full length PTX3 (Figure 1D).

**Figure 1.**
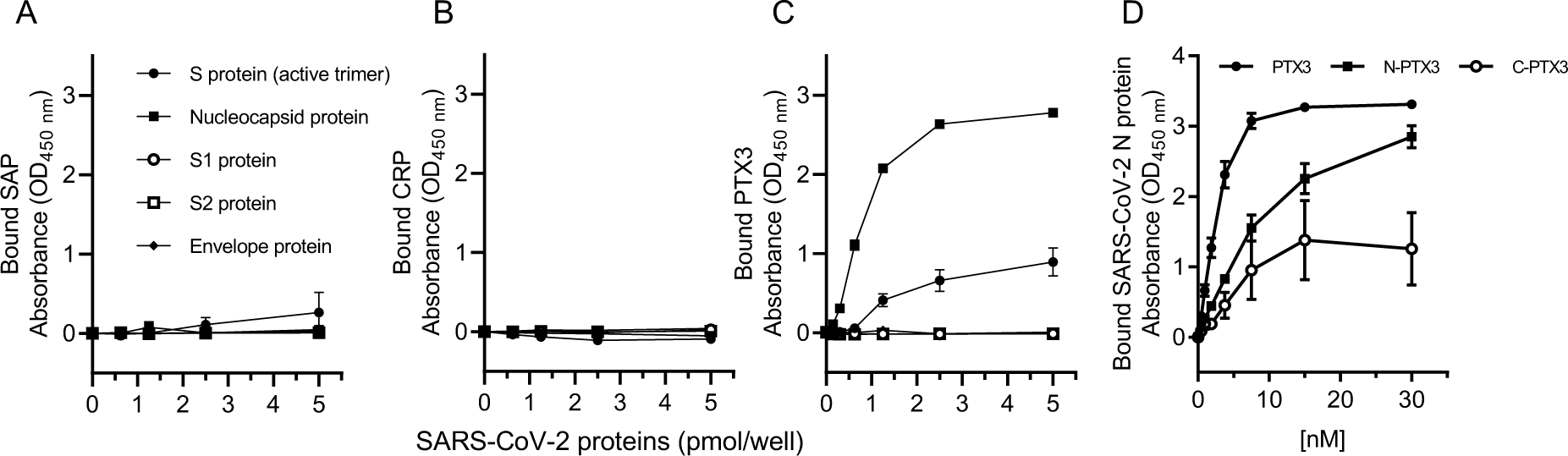
Interaction between pentraxins and SARS-CoV-2 proteins. (A-C) Recombinant HiS Tag SARS-CoV-2 proteins were immobilized on 96-well Nickel coated plates at different concentrations. Fixed concentrations of SAP (A), CRP (B) and PTX3 (C) were incubated over the captured viral proteins. Bound pentraxins were detected by ELISA with specific primary antibodies. (D) Full length PTX3 or its N- or C-terminal domains were captured on 96-well plates. Biotinylated SARS-CoV-2 Nucleocapsid protein was incubated at different concentrations. Bound nucleocapsid was detected by ELISA using HRP-conjugated streptavidin. All data are presented as mean ± SEM of two independent experiments performed in duplicate (n=4).

We next investigated the interaction between PRMs of the classical and the lectin pathway of complement (C1q and the collectin MBL, respectively) and the viral proteins. As shown in Figure 2A, C1q did not interact with any protein tested. In contrast, human MBL bound to SARS-CoV-2 Spike protein (Wuhan strain (Wu et al., 2020), active trimer), but not to the individual SARS-CoV-2 Spike subunits S1 [containing the receptor-binding domain (RBD)] and S2 (containing the membrane fusion domain) (Figure 2B). We validated these data by analyzing the binding of MBL to different recombinant SARS-CoV-2 Spike proteins obtained from different sources or produced in house either in HEK293 cells, or CHO cells, or in insect cells (Figure 2C and Extended Data Figure 1). All these preparations were bound by MBL, although with some differences. Notably, when we tested a non-covalent trimer of the SARS-CoV-2 Spike protein, we did not observe binding. These results indicate that a native- close structure of the SARS-CoV-2 Spike protein (presumably in the trimeric conformation) is indispensable for MBL recognition.

**Figure 2.**
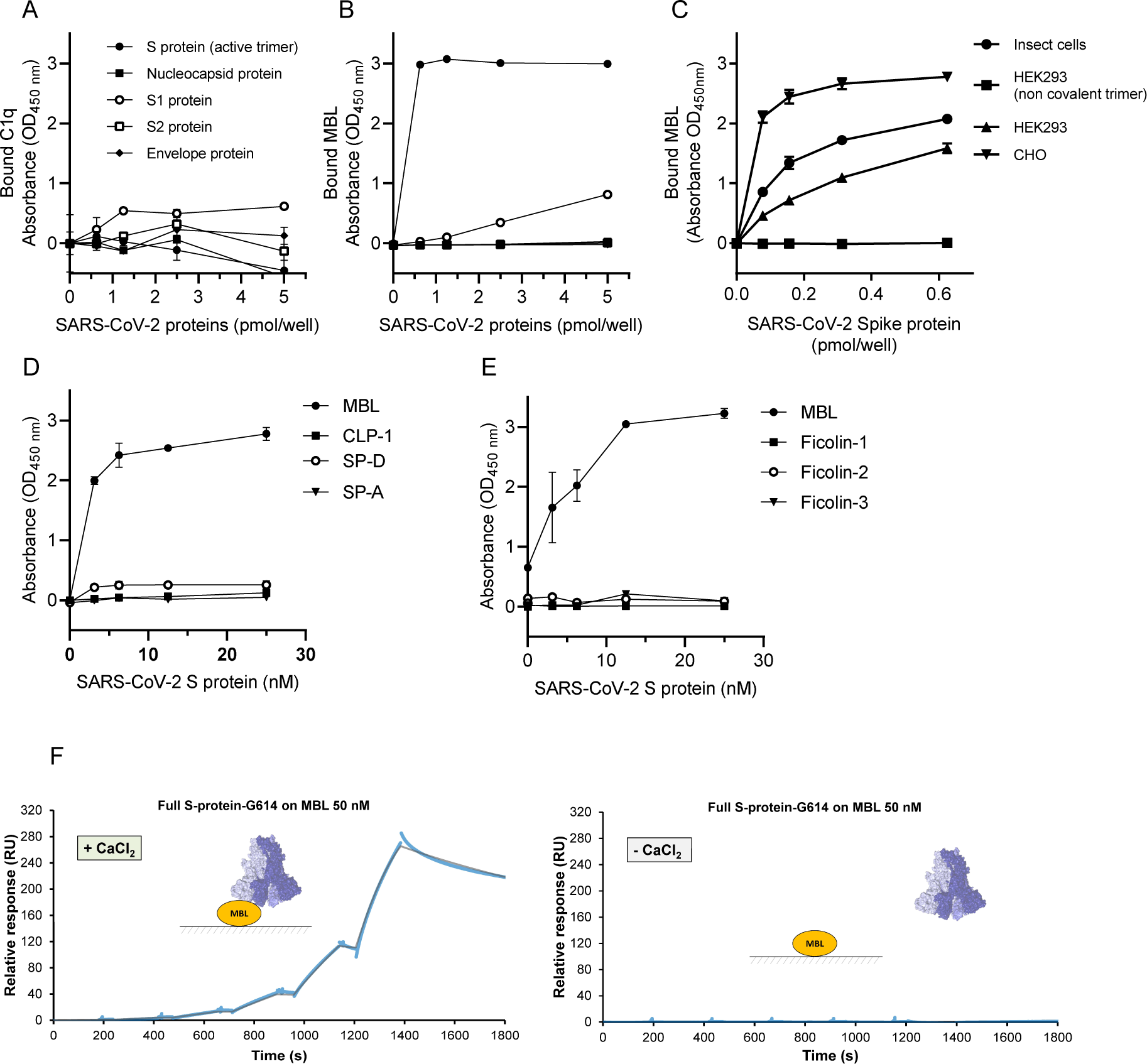
Interaction of C1q, MBL, ficolins and surfactant proteins with SARS-CoV-2 proteins. (A-C) Recombinant HiS Tag SARS-CoV-2 proteins were immobilized on 96-well Nickel coated plates at different concentrations. Fixed concentrations of C1q (A), or MBL (B) were incubated over the captured viral proteins. In (C), recombinant SARS-CoV-2 S proteins tested were expressed in different cell types. MBL (2 µg/mL- 6.7 nM) was incubated over the captured viral proteins. A-C: Bound proteins were detected by ELISA with specific primary antibodies. All data are presented as mean ± SEM of two independent experiments performed in duplicate (n=4). (D) MBL-, CLP-1-, SP-D-, SP-A- or (E) Ficolin-1-, Ficolin-2-and Ficolin-3-coated plates were incubated with various concentrations of biotinylated SARS- CoV-2 S protein. D-E: Bound S protein was detected by ELISA with HRP-conjugated streptavidin (mean ± SEM of two independent experiments in duplicate, n=4). (F) SPR shows binding of recombinant full Spike protein trimer to immobilized MBL (KD=34 nM, left). No binding is detected in the absence of CaCl2 (right).

MBL is a member of the collectin family, a class of PRMs composed of a Ca^2+^- type lectin domain (also called Carbohydrate Recognition Domain, CRD) and a collagen-like domain (Holmskov et al., 2003). Thus, we analyzed the interaction of SARS-CoV-2 Spike protein with other collectins involved in innate immunity, such as Collectin-12 (CLP-1) and the pulmonary surfactant proteins SP-A and SP-D. We also extended the analysis to recombinant Ficolin-1, -2, or -3, a family of proteins known to activate the complement lectin pathway, and structurally-related to MBL. As shown in Figure 2D and 2E, in contrast with MBL, CLP-1, SP-A, SP-D, and ficolins did not bind to SARS-CoV-2 Spike protein, indicating that recognition of Spike is unique to MBL.

We further characterized the interaction of SARS-CoV-2 Spike protein with MBL by Surface Plasmon Resonance (SPR). Different concentrations of recombinant, SARS-CoV-2 Spike protein or RBD domain were flowed onto MBL immobilized on the biosensor surface. As shown in Figure 2F and Extended Data Figure 2, trimeric SARS-CoV-2 Spike protein formed a stable calcium-dependent complex with nanomolar affinity (K_D_=34 nM) whereas MBL did not bind the isolated RBD, confirming the results obtained using the S1 subunit.

To mimic the interaction between MBL and SARS-CoV-2 Spike protein in its physiological conformation in the viral envelope, we investigated the binding of viral particles of SARS-CoV-2 Spike protein pseudotyped on a lentivirus vector to MBL-coated plates. The interaction was determined by lysing the bound pseudovirus and measuring the lentiviral vector p24 core protein by ELISA. While control lentiviral particles pseudotyped with the VSV-g glyprotein (VSV-pseudovirus) did not result in any binding, those exposing the SARS-CoV-2 Spike protein showed a specific interaction with MBL (Figure 3A). These data strongly suggest that MBL can also interact with the SARS-CoV-2 Spike protein exposed on the virus surface.

**Figure 3.**
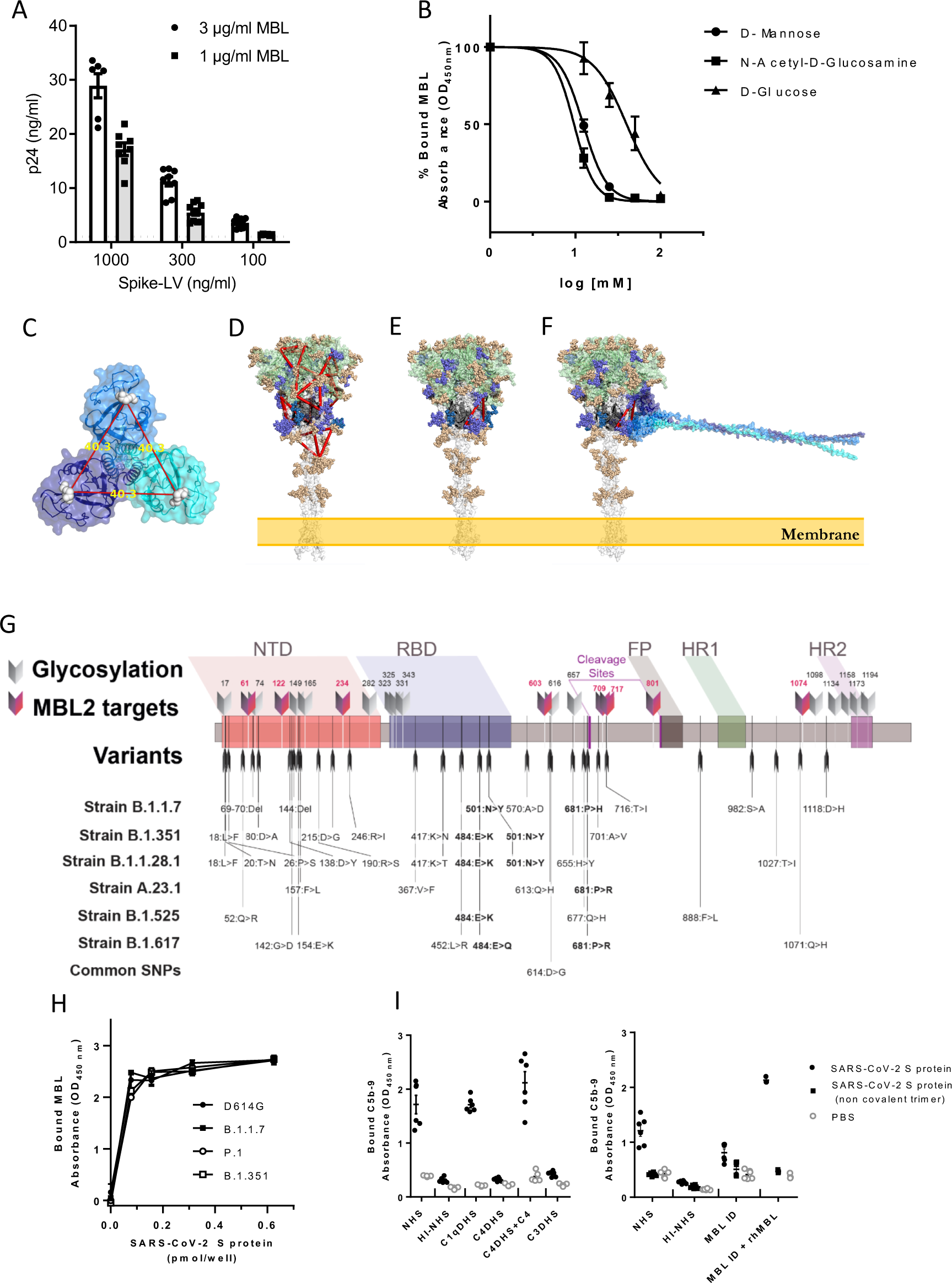
Interaction of MBL with SARS-CoV-2 S protein through its Carbohydrate Recognition Domain (CRD). (A) MBL-coated plates were incubated with different concentrations of viral particles of SARS-CoV-2 Spike protein (Spike-LV) or VSV-g pseudotyped on a lentivirus vector. After lysis, bound pseudovirus was quantified by measuring the released p24 core protein by ELISA. Dotted line represents p24 levels measured using the highest concentration of VSV-g pseudotyped lentivirus. Data are represented as mean ± SEM of two independent experiments (n=6-10). (B) Biotinylated SARS-CoV-2 S protein was captured on Neutravidin coated plates. MBL (0.25 μg/mL) was incubated over S protein, alone or in the presence of different concentrations of D-mannose or N-acetyl-glucosamine or D-glucose. Binding of free MBL was detected by ELISA. Data are presented as percentage of bound MBL (mean ± SEM of two independent experiments, n=4). (C) Trimeric MBL2 model shows the distance of approximately 40 Å between mannose-binding sites. MBL2 subunits are represented in transparent surface and cartoon, colored in cyan, blue, and dark blue; mannose molecules are represented in white spheres; (D) 14 mannose-binding sites, represented as red triangles, imposed onto Spike protein. Spike is represented in transparent surface and cartoon, where S1 region (1-685) is colored in green, beginning of S2 region (686-815) is colored in black, and S2’ region is colored in white. (E) Putative binding site of MBL2. Pose with the highest site-specific probability to be glycosylated with oligomannose; (F) Spike-MBL2 complex. Glycosylation sites are colored according to the oligomannose glycosylation probability. Gold < 60%. Purple > 80% up until S2’ region. Blue > 80% in the S2’ region. (G) Schematic representation of glycosylation sites and nucleotide substitutions in the variant strains identified to date. The 8 glycosylation sites containing oligomannose-types glycans, which are potential targets of MBL, are evidenced. SNPs common to all variants are in bold. NTD: N terminal domain; RBD: receptor binding domain; FP: fusion peptide; HR1 and 2: heptad repeat 1 and 2; cleavage sites are reported. (H) Binding of MBL to SARS-CoV-2 S protein variants: recombinant SARS-CoV-2 S protein (D614G), B.1.1.7, P.1, and B.1.351 variants were captured onto a Nickel coated plate at different concentrations. MBL (1 µg/mL – 3.4 nM) was incubated over the captured viral proteins. Data are represented as mean ± SEM, n=3 replicates. One representative experiment out of two performed is shown for B.1.1.7. For the other variants, one experiment was performed. (I) MAC (C5b-9) deposition on SARS-CoV- 2 Spike protein-coated plates. The assay was performed in the presence of 10% normal human serum (NHS), C1qDHS, C4DHS, C4DHS reconstituted with C4, C3DHS, MBL-ID serum, MBL-ID serum reconstituted with rhMBL. Mean ± SEM of two independent experiments, performed in triplicate (n=6).

### MBL interacts with glycosidic sites of the SARS-CoV-2 spike protein

The SARS-CoV-2 Spike protein is highly glycosylated, as recently described (Watanabe et al., 2020). Out of the 22 N-glycosylation sites, 8 contain oligomannose-types glycans, which could be interaction sites for the MBL carbohydrate recognition domain (CRD).

To address this possibility, we performed a solution-based competition assay with D-mannose and N-acetyl-glucosamine, two specific ligands of the lectin. As shown in Figure 3B, D-mannose and N-acetyl-glucosamine inhibited MBL binding to the Spike protein, thus confirming the Ca^2+^-dependent interaction between the MBL lectin domain and the glycosidic sites exposed by the Spike protein. D-Glucose, a non-specific ligand of MBL, inhibited the interaction only at higher concentration (Figure 3B). Based on the alignment of MBL crystal structure with mannose molecules (Fig. 3C), we identified 14 putative binding sites on the Spike protein (Fig. 3D). Next, we considered sites having a high (>80%) oligomannosylation occupancy (Watanabe et al., 2020). This analysis provided two possible MBL binding sites, namely N603, N801 and N1074 all on the same Spike chain, or N603, N1074 and N709 with N709 on a neighboring chain (Figure 3E). Interestingly, in both cases, the hypothesized MBL binding sites spans across the S1 and S2 region (Figure 3F) of the Spike protein providing hints to a possible inhibition mechanism. These data indicate that the glycosylation state of the SARS-CoV-2 Spike protein is important for its interaction with MBL.

### Interaction of MBL with Spike from VoC

We then tested whether MBL recognized Spike proteins from VoC. First, we analyzed whether the known 22 glycosylation sites of each protomer are affected by the reported mutations. Figure 3G shows a schematic representation of the 22 positions of N-linked glycosylation sequons and of 35 known mutations, indicating that none of these mutations involve the glycosylation sites, and suggesting that MBL could interact with the variants with the same affinity. In agreement with our binding assays, no MBL target sites are expected in the RBD. We assessed by solid phase assay the interaction of MBL with the SARS-CoV-2 D614G Spike trimeric protein, the B.1.1.7 variant (emerged in UK), the B.1.1.28 or P.1 variant (emerged in Brazil), and B.1.351 (emerged in South Africa) (Figure 3H). In agreement with the *in silico* analysis, MBL bound the VoC Spike proteins tested with similar affinity.

### Complement lectin pathway activation

We asked whether the interaction of MBL with Spike could activate the complement lectin pathway. We incubated SARS-CoV-2 Spike protein-coated plates with human serum, or C1q- or C4- or C3-depleted serum, and we assessed the deposition of C5b-9. As shown in Figure 3I (left panel), incubation with either normal human serum or C1q-depleted serum resulted in complement deposition mediated by SARS-CoV-2 Spike protein. Conversely, incubation with a serum depleted of C4 strongly reduced C5b-9 deposition, with levels comparable to those observed with heat-inactivated serum or C3-depleted serum. Reconstitution of C4-depleted serum with purified C4 restored C5b-9 deposition levels similar to those observed with normal human serum. To further address the role of MBL in SARS-CoV-2 Spike protein-mediated complement activation, we assessed C5b-9 deposition by incubating normal human serum or MBL-immunodepleted serum over captured SARS-CoV-2 Spike protein, either as active, or non-covalent trimer (Figure 3I, right panel). In agreement with binding data, no complement deposition was observed with the non-covalent trimeric Spike protein. Notably, immunodepletion of MBL from human serum resulted in a significant reduction in C5b-9 deposition, which could be fully reverted by addition of rhMBL (Figure 3I, right panel). These data clearly indicate that SARS-CoV-2 Spike, by interacting with MBL, activates the complement lectin pathway.

### SARS-CoV-2 inhibition by MBL

To validate the relevance of the interaction between MBL and SARS-CoV-2 Spike protein, we investigate whether MBL inhibited SARS-CoV-2 entry in susceptible cells. We first tested the effect of MBL and other soluble PRMs (10-fold serial dilution, from 0.01 to 10 µg/ml) on the entry of the viral particles of SARS-CoV-2 Spike protein pseudotyped on a lentivirus vector in 293T cells overexpressing Angiotensin-Converting Enzyme 2 (ACE2). Among the soluble PRMs tested, MBL was found to be the only molecule with anti-SARS-CoV-2 activity. Spike-mediated viral entry was inhibited by 90% at the highest concentration of 10 µg/ml (34 nM) with an EC50 value of approximately 0.5 µg/ml (1.7 nM) (Figure 4A). As control, entry of lentiviral particles pseudotyped with the VSV-g glycoprotein was not inhibited by MBL (Figure 4A).

**Figure 4.**
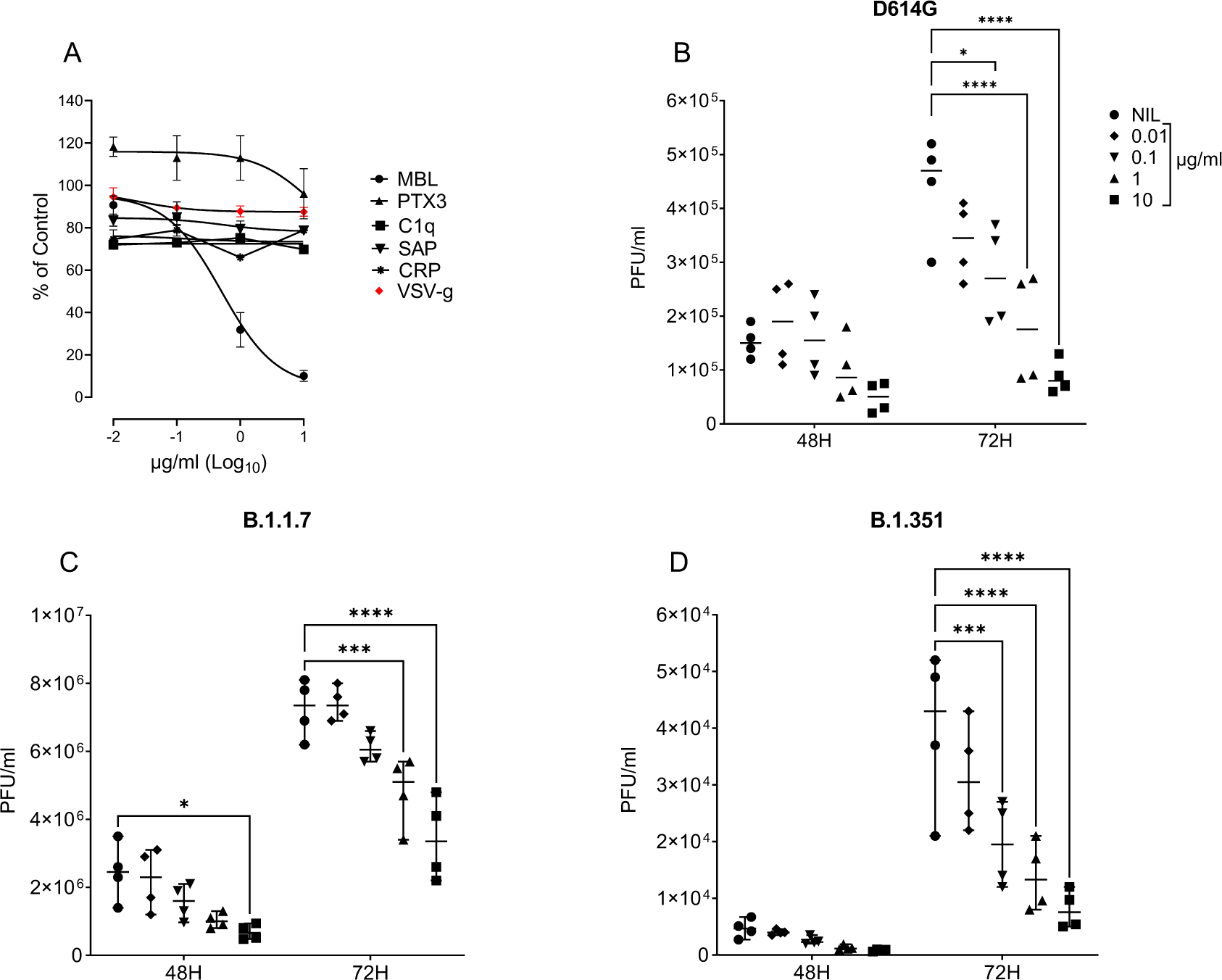
Inhibition of SARS-CoV-2 infection by MBL. (A) Entry of lentiviral particles pseudotyped with SARS-CoV-2 Spike protein in 293T cells overexpressing ACE2 in the presence of ten-fold serial dilutions of humoral innate immunity PRMs (from 0.01 to 10 µg/ml). As control, entry of lentiviral particles pseudotyped with the VSV-g glycoprotein was tested in parallel in the presence of MBL. Percentage of control was calculated as ratio of GFP-positive cells in the presence of humoral PRMs and the GFF-positive cells in the absence of humoral PRMs. Data are means ± SEM of three independent experiments in triplicates with the curves representing a three-parameter dose response model. (B-D) Inhibition of the infectivity of the D614G (isolate EPI_ISL_413489) (B), B.1.1.7 (C) and B.1.351 (D) SARS-CoV-2 variants by MBL in Calu-3 cells. SARS-CoV-2 (MOI=1) and Calu-3 cells were preincubated in complete medium containing different concentrations of MBL (0.01–10 µg/mL– 0.034-34 nM) before infection. After 48 and 72 h, the infectivity of SARS-CoV-2 present in cell culture supernatants was determined by a plaque-forming assay in Vero cells. NIL: no MBL. Mean values of two (B) or one (C, D) experiments in duplicate cell culture are shown. ****p value <0.0001, ***p value <0.001, **p value <0.01 as determined by two-way ANOVA with Bonferroni’s correction.

We next tested the antiviral activity of MBL on the SARS-CoV-2 infection of lung epithelial models relevant to human infections. Among a number of lung-derived epithelial cell lines, Calu-3 (human lung adenocarcinoma) cells have been shown to be permissive to SARS-CoV-2 infection (Chu et al., 2020). SARS-CoV-2 (D614G variant, MOI=0.1 and 1) was preincubated in complete medium containing different concentrations of MBL (0.01–10 µg/mL; 0.034-34 nM) before incubation with Calu-3 cells. After 48 and 72 h, the infectivity of SARS-CoV-2 present in cell culture supernatants was determined by a plaque-forming assay in monkey-derived Vero cells. Vero cells are a handy cell line used worldwide as it is devoid of the interferon (IFN) response (Desmyter et al., 1968) and, for this reason, highly supportive of virus replication.

As shown in Extended Data Figure 3A, MBL showed a concentration-dependent inhibition of SARS-CoV-2 infection of Calu-3 cells at MOI 0.1 (upper panel) and 1 (lower panel), that was statistically significant at 1 and 10 µg/ml (3.4 and 34 nM) 72 h after infection. When both virus and cells were pre-incubated with the same concentrations of MBL (0.01–10 µg/mL; 0.034-34 nM), the antiviral activity increased significantly from 0.1 µg/ml (0.34 nM) to the top concentration of 10 µg/ml (34 nM), 72 h post-infection (PI) (Figure 4B and Extended Data Figure 3B). The calculated EC50 was 0.08 µg/mL (0.27 nM) at 72h. Notably, MBL showed a concentration-dependent inhibition of infection of Calu-3 cells also by SARS-CoV-2 variant 20I/501Y.V1 (B.1.1.7) at MOI 1 (Figure 4C) and MOI 0.1 (Extended Data Figure 3C), as well as by 20H/501Y.V2 (B.1.351) at MOI 1 (Figure 4D).

Furthermore, a model of 3D-human bronchial epithelial cells (HBEC) was used to test whether MBL inhibited SARS-CoV-2 replication. SARS-CoV-2 production at the epithelial apical surface increased sharply at 48 h PI (not shown), reaching 48x10^6^ ± 6x10^6^ (mean ± SEM) PFU/ml 72 h PI. Treatment of HBEC with MBL decreased viral production to 4x10^6^ ± 0.8x10^6^ PFU/ml 72 h PI at the highest concentration of 50 µg/ml (170 nM) (Figure 5A). In contrast PTX3 treatment was ineffective at inhibiting virus production (Extended Data Figure 3D). We then assessed whether in these experimental conditions, MBL affected inflammatory responses in HBEC upon SARS-CoV-2 infection. As shown in Extended Data Figure 3E, MBL treatment inhibited the production of IL-8 and CXCL5, chemokines involved in myeloid cell recruitment and activation.

**Figure 5.**
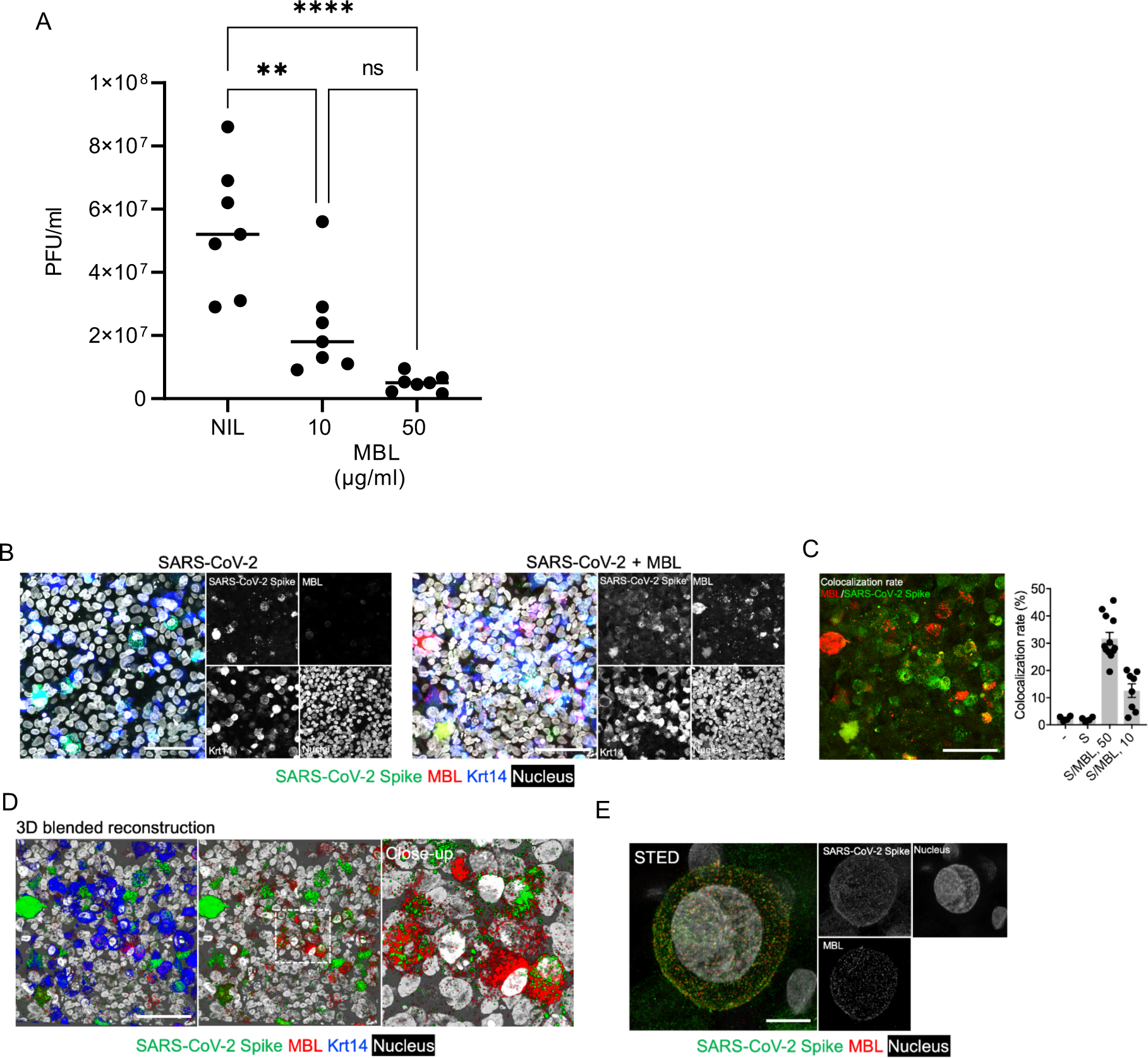
Inhibition by MBL of SARS-CoV-2 infection of primary respiratory cells. (A) SARS-CoV-2 production at the HBEC apical surface at 72 h PI, in the presence of 10 or 50 µg/ml (34 or 170 nM) MBL. Mean values of three experiments in triplicate (2 donors) or single (1 donor) cell cultures are shown. A: p values were determined by one-way ANOVA with Bonferroni’s correction. ****: p value <0.0001, **: p value <0.01. (B-E) Colocalization of SARS-CoV-2 S protein and MBL in infected HBEC. (B), Confocal analysis of the localization of SARS-CoV-2 S protein (green), MBL (red) in HBEC cultures infected by SARS-CoV-2 in presence or not of MBL (50 µg/ml -170 nM). Left panels, merged images of fluorescence signals; right panels, single signals extracted. Representative MIP images of 4-12 *Z-* stacks acquired in tiling modality are shown. Bar, 30µm. (C) Left panel: extracted signals of SARS-CoV-2 S protein and MBL of (B). Right panel: colocalization rate between SARS-CoV-2 S protein and MBL (10 or 50 µg/ml - 34 or 170 nM). Each spot corresponds to a single *XYZ* image presented as MIP. Mean±SE. (D) 3D rendering of B right panel, showing a blended reconstruction of the localization between SARS-CoV-2 S protein and MBL in HBEC cultures. Left panel, contribution of merged signals. Bar, 30µm. Middle panel, extracted image of signal of SARS-CoV-2 S protein and MBL. Right panel, close-up image that refers to the area dashed in white. (E) STED analysis of the localization of SARS-CoV-2 S protein and MBL in HBEC. Left panel, merged signals of SARS-CoV-2 S protein and MBL and nucleus; right, single signals extracted. Bar, 3µm.

We finally evaluated occurrence of MBL-Spike protein interaction in SARS-CoV-2 infected HBEC by confocal microscopy. As shown in Figure 5B and C, MBL colocalized with SARS-CoV-2 Spike protein in infected cells. In 3D rendered images of the HBEC cell cultures (Figure 5D and Movie S1), colocalization was preferentially associated to the apical side of cytokeratin 14 positive cells. Evidence of the interaction between MBL and SARS-CoV-2 Spike protein in infected HBEC at molecular scale (<100nm *XY* spatial resolution) were also obtained in STED-based super-resolution microscopy (Figure 5E).

### *MBL2* haplotypes are associated with severe COVID-19

MBL2 genetic variants have been shown to correlate with increased susceptibility to selected infections, including SARS (Ip et al., 2005). To explore the significance of our *in vitro* results in the frame of COVID-19 pandemic, we investigated the possible association of *MBL2* polymorphisms with severe COVID-19 with respiratory failure in an Italian cohort of 332 cases and 1,668 controls (general population). We initially focused on six SNPs known to be associated with MBL2 protein levels (Table 1) (Lipscombe et al., 1992; Madsen et al., 1994; Madsen et al., 1995; Sumiya et al., 1991). Surprisingly, we observed a significant difference only in the frequency of the rs5030737-A allele between patients and controls (7.7% and 6.0%, respectively; OR=1.43, 95%CI=1.00-2.05, P=0.049; Table 1A), which however did not survive the correction for multiple testing. When we compared the frequencies of haplotypes determined by all six SNPs, we found the CCGGCC haplotype frequency significantly decreased in patients with severe COVID-19 (26.7% in cases, 30.4% in controls). This haplotype shows a protective effect (odds ratio (OR)=0.78, 95%CI=0.65-0.95, P=0.025; Table 1B), consistently with the lack of the rs5030737-A allele, which is only present in the CCAGCC haplotype (OR=1.38, 95%CI=1.00-1.90; P=0.078; Table 1B).

**Table 1.**
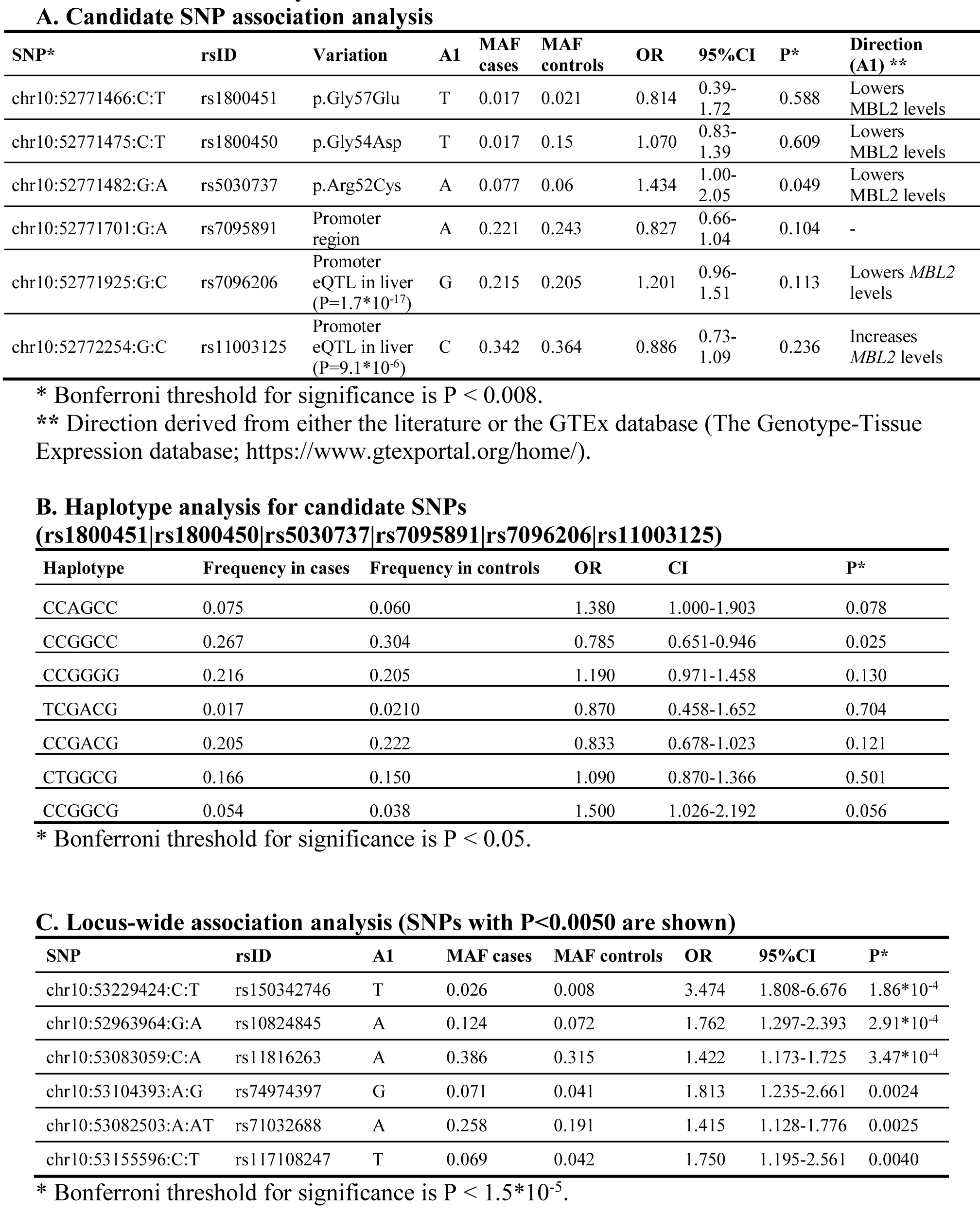
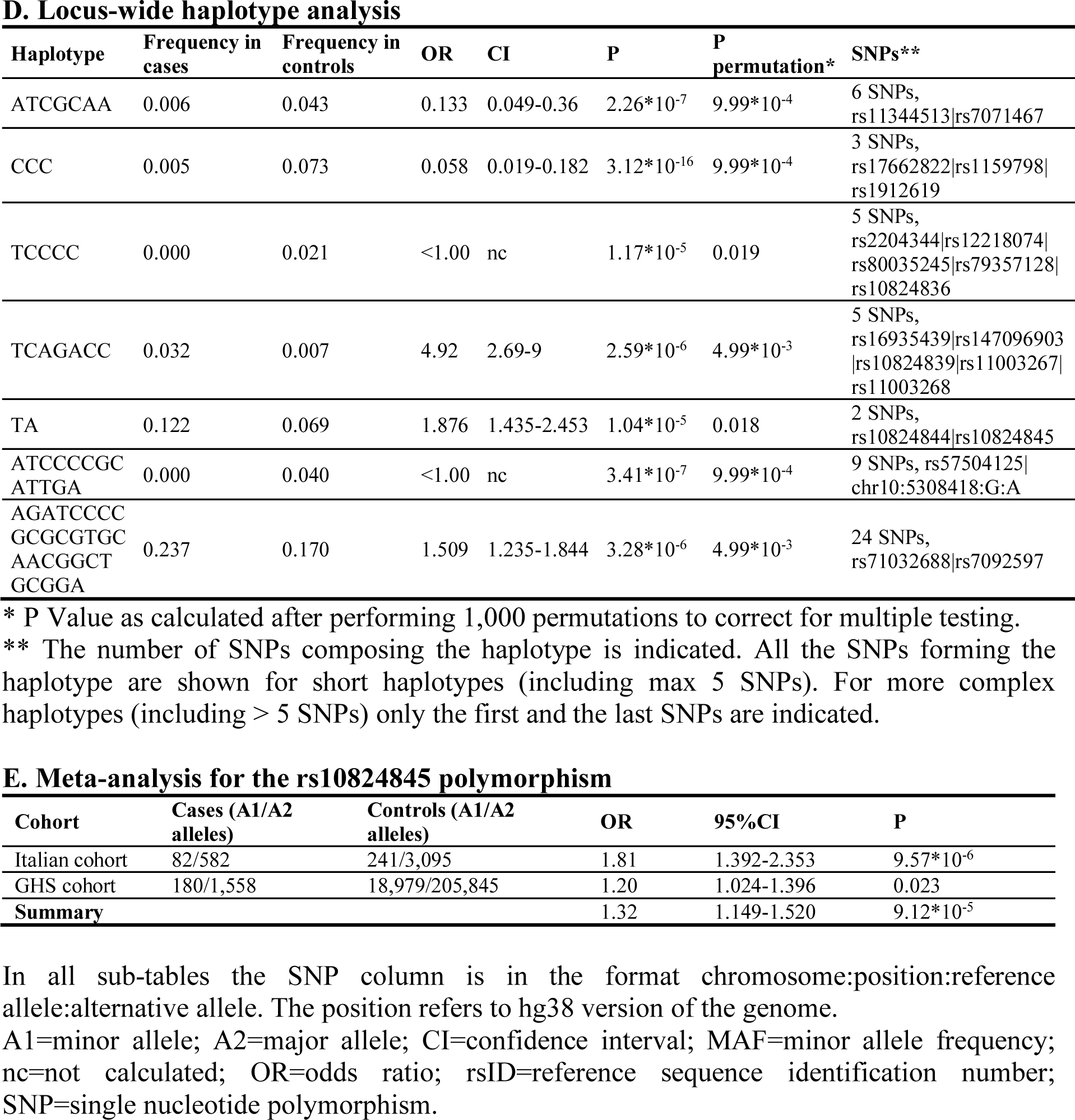
Association analysis results

Though borderline, these first association results encouraged us to investigate the 1- Mb-long genomic region encompassing the *MBL2* gene systematically. To this aim, we performed single-SNP as well as haplotype-based association analyses using genotyped/imputed data on 3,425 polymorphisms. Single-SNP association analysis revealed three suggestive signals (rs150342746, OR=3.47, 95%CI=1.81-6.68, P=1.86*10^-4^; rs10824845, OR=1.76, 95%CI=1.30-2.39, P=2.91*10^-4^; and rs11816263, OR=1.42, 95%CI=1.17-1.73, P=3.47*10^-4^; Table 1C; Figure 6), whereas haplotype-based analysis disclosed 7 haplotypes of different lengths, from 2 to 24 SNPs, strongly associated with severe COVID-19 (all surviving the correction for multiple tests; Figure 6; Table 1D). Among them, the one composed of polymorphisms rs10824844-rs10824845 incorporates one of the two top-markers evidenced by the single-SNP association analysis and is present in 12.2% of cases and 6.9% of controls (TA haplotype, OR=1.88, 95%CI=1.44-2.45, P=1.04*10^-5^; Table 1D). Hence, we performed a meta-analysis based on the rs10824845 polymorphism by including the GHS study of COVID-19 patients: this resulted in a pooled OR=1.32, 95%CI=1.15-1.52, P=9.12*10^-5^ (Table 1E). Notably, the rs10824845 polymorphism points to a regulatory region characterized by the presence of an enhancer (GH10J052964), described as a distant modulator of the *MBL2* gene, active in HepG2 cells (hepatocytes), as well as M0 (from venous blood) and M1 (from cord and venous blood) macrophages (data from the GeneHancer database, available through http://www.genecards.org/).

**Figure 6:**
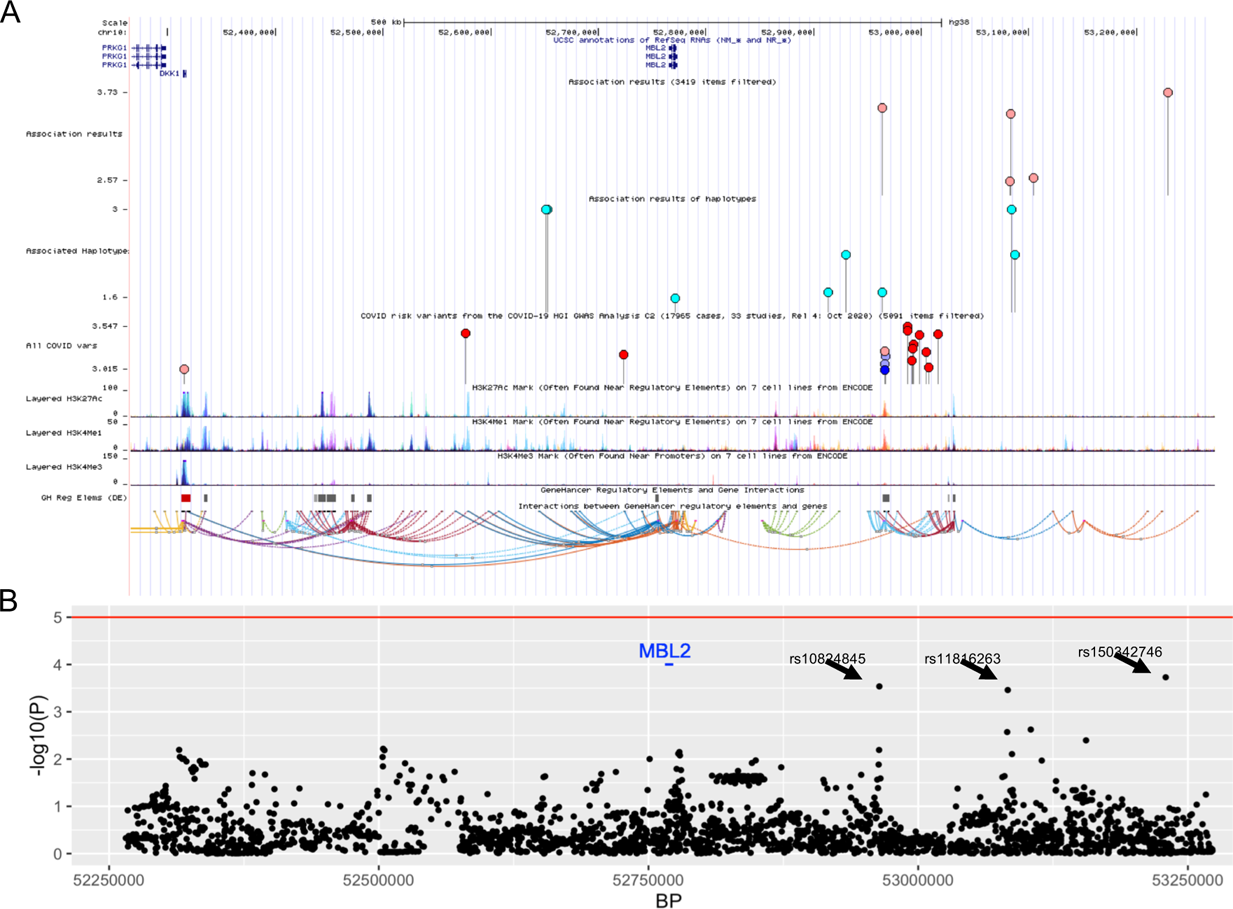
The *MBL2* locus: structure and main association signals with severe COVID- 19. A screenshot from the UCSC Genome browser (http://genome.ucsc.edu/; release Dec. 2013, GRCh38/hg38) specifically highlighting the 1-Mb region surrounding the *MBL2* gene is shown. The panel reports, in order, the following tracks: i) the ruler with the scale at the genomic level; ii) chromosome 10 nucleotide numbering; iii) the UCSC RefSeq track; iv) COVID-19 risk variants from our study (lollipops show only signals at P<3*10^-3^); v) COVID- 19 risk haplotypes, marked by the tagging SNP, from our study (lollipops show all haplotypes reported in Table 1B and 1D); vi) COVID-19 risk variants from the COVID-19 HGI GWAS Analysis C2 (17,965 cases, 33 studies, Release 4: October 2020); vii) ENCODE data (https://www.encodeproject.org/) for H3K27Ac, H3K4Me1, H3K4Me3 histone modifications marks, all derived from 7 cell lines; viii) the GeneHancer regulatory elements track. (B) The Manhattan plot of the single-SNP association analysis is reported. The horizontal line represents the suggestive P=5*10^−5^ significance level. SNPs showing lowest P value signals are indicated by an arrow. Bonferroni threshold for significance corresponds to P < 1.5*10^-5^.

## Discussion

Among the 10 fluid phase PRM tested in this study, only PTX3 and MBL bound SARS-CoV-2 virus components. PTX3 recognized the viral Nucleoprotein and had no antiviral activity. PTX3 was expressed at high levels by myeloid cells in blood and lungs and its plasma levels have strong and independent prognostic significance for death in COVID-19 patients (Brunetta et al., 2021; Schirinzi et al., 2021). It remains to be elucidated whether PTX3 plays a role in Nucleocapsid-mediated complement activation and cytokine production (Gao et al., 2020; Karwaciak et al., 2021; McBride et al., 2014).

MBL recognized the SARS-CoV-2 Spike protein, including that of three VoC, and had antiviral activity *in vitro*. MBL had previously been shown to bind SARS-CoV Spike (Zhou et al., 2010). The interaction of MBL with Spike required a trimeric conformation of the viral protein, did not involve direct recognition of the RBD and was glycan-dependent, as expected. Site-specific glycosylation analysis of the SARS-CoV-2 Spike protein revealed the presence of various oligomannose-type glycans across the protein (Watanabe et al., 2020).

Molecular modelling reported here suggests that the MBL trimer interact with glycans attached to the residues N603, N801 and N1074 on the same chain or N603, N709 and N1074 with N709 on a different chain. In both cases the hypothesized MBL binding site spans across the S1 and S2 region of SARS-CoV-2 Spike, suggesting a possible neutralization mechanism.

The binding of MBL could prevent the detachment of the S1 region and the release of the fusion peptide at position 815, thus inhibiting virus entry into host cells. However, the mechanisms responsible for the antiviral activity of MBL remain to be fully defined. It is noteworthy that C-type lectins have been reported to act as entry receptors (or coreceptors) (Chiodo et al., 2020; Lempp et al., 2021; Lu et al., 2021) and MBL is likely to compete at this level.

Interestingly, the *in silico* analysis presented here indicate that mutations in variants reported until now do not affect glycosylation sites containing oligomannose-types glycans potentially recognized by MBL. In addition, binding and infection experiments show that the anti-viral activity of MBL is not affected by these mutations. This indicate that the glycosylation sites are generally spared by selective pressure, suggesting they are essential for SARS-CoV-2 infectivity. It has been recently shown that mechanisms of *in vitro* escape of SARS-CoV-2 from a highly neutralizing COVID-19 convalescent plasma include the insertion of a new glycan sequon in the N-terminal domain of the Spike protein, which leads to complete resistance to neutralization (Andreano et al., 2020). This result further emphasizes the relevance of Spike glycosidic moieties targeted by MBL in SARS-CoV-2 infectivity.

MBL was found to interact with Spike and have antiviral activity with an EC50 of approximately 0.08 μg/ml (0.27 nM) and an affinity of 34 nM. These concentrations are well in the range of those found in the blood of normal individuals (up to 10 μg/ml) which increase 2-3-fold during the acute phase response. MBL plasma levels in healthy individuals are extremely variable, in part depending on genetic variation in the *MBL2* gene. Defective MBL production has been associated with increased risk of infections, in particular in primary or secondary immunodeficient children (Koch et al., 2001). In SARS, conflicting results have been reported concerning the relevance of *MBL2* genetic variants in this condition (Ip et al., 2005; Yuan et al., 2005; Zhang et al., 2005). In COVID-19, one *MBL2* SNP has been associated with development and severity of the infection (Medetalibeyoglu et al., 2021). We investigated the possible role of *MBL2* genetic variants in determining susceptibility to severe COVID-19 with respiratory failure, by exploiting the statistical power provided by haplotype analysis. Surprisingly and in contrast with a previous study (Medetalibeyoglu et al., 2021), we found only a borderline correlation between one haplotype of the 6 SNPs associated with MBL levels and frequency of severe COVID-19 cases. However, we found a total of 7 significantly associated haplotypes, distributed along the *MBL2* genomic region, often mapping in correspondence of regulatory elements (such as enhancers, promoter region, histone marks; Figure 6). Our association data are reinforced by: i) the meta-analysis results, obtained by integrating the summary statistics from a European cohort of >113,000 individuals; ii) the fact that one of our second best associations (i.e., rs10824845) maps in proximity of a cluster of suggestive signals (P<5*10^-4^) identified by the COVID-19 Host Genetic Initiative (https://www.covid19hg.org/; results on release 4 include data from up to 33 different worldwide studies; Figure 6); iii) the Regeneron – Genetic Center database, which contains association results also for rare variants (MAF<1%), reports a significant association (P<0.05) for the rs35668665 polymorphism both with susceptibility to COVID-19 (OR=4.11, GHS cohort) and with severity of symptoms (OR=7.91, UK BioBank cohort). Interestingly, this variant maps in correspondence of the last nucleotide of *MBL2* exon 1, thus possibly interfering with the splicing process; and iv) the rs5030737 (p.Arg52Cys) polymorphism in *MBL2* has been described in the UKBiobank ICD PheWeb database (https://pheweb.org/UKB-SAIGE/) as a top signal in determining both “dependence on respirator [Ventilator] or supplemental oxygen” (ICD code Z99.1; P=2.7*10^-4^) and “Respiratory failure, insufficiency, arrest” (ICD code J96; P=2.7*10^-3^). These observations suggest that genetic variations in *MBL2*, possibly involved in the modulation of the expression of the gene in hepatocytes, and, interestingly, in macrophages, could play a role in determining susceptibility to severe COVID-19 with respiratory failure. Therefore genetic analysis is consistent with the view that MBL recognition of SARS-CoV-2 plays an important role in COVID-19 pathogenesis.

Upon interaction with Spike, MBL was found to activate the lectin pathway of complement, as expected. Complement has been credited an important role in the hyperinflammation underlying severe disease and is considered a relevant therapeutic target (Carvelli et al., 2020; Risitano et al., 2020). Therefore, as for innate immunity in general including the IFN pathway (King and Sprent, 2021), MBL-mediated recognition of SARS-CoV-2 may act as a double-edged sword. In early phases of the disease MBL, possibly produced locally by macrophages, may serve as a mechanism of antiviral resistance by blocking viral entry, whereas in advanced disease stages it may contribute to complement activation and uncontrolled inflammation.

MBL has been safely administered to patients with cystic fibrosis and chronic lung infections in which MBL deficiency contributes to pathogenesis (Garred et al., 2002; Jensenius et al., 2003). Therefore, the results presented here have translational implications both in terms of comprehensive genetic risk assessment and development of local or systemic therapeutic approaches.

## Data Availability

The data that support the findings of this study are available from the corresponding authors upon request.

## Acknowledgements

This work was supported by a philanthropic donation by Dolce & Gabbana fashion house (to A.M., C.G., E.V.), by the Italian Ministry of Health for COVID-19 (to A.M. and C.G.), the Italian Ministry of University and Research (to P.I.), by the Department of Excellence project PREMIA (PREcision MedIcine Approach: bringing biomarker research to clinic, to P.I.).We also thank the generous contribution of Banca Intesa San Paolo, and AMAF Monza ONLUS and AIRCS for the unrestricted research funding.

## Author contributions

A.M. and C.G. conceived the study in March 2021 and catalyzed the interaction between different Institutions; C.G., E.V. and B.B. supervised the development of the effort; M.S. and I.P. conducted the core experimental work related to binding, antiviral activity and Complement activation. The genetic analysis was conducted by E.M.P. supervised by S.D. and R.A.; A.D. performed the imaging analysis; M.P., L.V., M.M and A.C. performed SPR analysis and modeling; S.N.M. performed the bioinformatic analysis; M.G.U., R.R., P.I. provided essential tools and material; F.S., Ma.S., D.C., P.G., N.P., V.C. N.C., N.M. conducted complementary experiments.

## Methods

### Recombinant proteins and Antibodies

Commercial and non-commercial recombinant SARS-CoV-2 proteins used in this study are listed in Supplementary Table 1. Recombinant human PTX3 and its domains from CHO cells were produced in house, as previously described (Bottazzi et al., 1997). Recombinant human Surfactant Protein-A (SP-A) was from Origene. Recombinant human MBL, Collectin-12 (CLP-1), Ficolin-1, Ficolin-2, Ficolin-3 and Surfactant Protein-D (SP-D) were from Biotechne. Purified human C1q was from Complement Technology, purified CRP was from Millipore and purified SAP was purchased from Abcam. Rabbit anti-PTX3 antibody was produced in house (Bottazzi et al., 1997), rabbit anti-MBL Ab was purchased from Abcam. Anti-C1q polyclonal antibody was purchased from Dako. Anti-CRP and anti-SAP antibodies were from Merck.

### Binding of Humoral Pattern Recognition Molecules to SARS-CoV-2 proteins

Recombinant His-Tag SARS-CoV-2 proteins were immobilized at different concentrations (ranging from 6.25 to 50 pmol/mL) on 96-well Nickel coated plates (Thermo Fisher Scientific) for 1 h at room temperature. Plates were then blocked for 2 h at 37°C with 200 µL of 2% BSA diluted in 10 mM Tris-HCl buffer, pH 7.5, containing 150 mM NaCl, 2 mM CaCl2 and 0.1% Tween-20 (TBST-Ca^2+^). Following blocking, plates were washed three times with TBST-Ca^2+^ and incubated for 1 h at 37°C with 100 µL PTX3 (4 µg/mL – 12 nM in TBST-Ca^2+^), MBL (1-2 µg/mL – 3.4-6.7 nM in TBST-Ca^2+^), C1q (4 µg/mL – 10 nM in TBST-Ca^2+^), CRP (3 µg/mL – 25 nM in TBST-Ca^2+^) and SAP (4 µg/mL – 32 nM in TBST-Ca^2+^). After washes, plates were incubated for 1 h at 37°C with specific primary antibodies, followed by the corresponding HRP-conjugated secondary antibodies. Both primary and secondary antibodies were diluted in TBST-Ca^2+^ buffer. After development with the chromogenic substrate 3,3’,5,5’-tetramethylbenzidine (TMB, Thermo Fisher Scientific, USA), binding was detected by absorbance reading at 450 nm on a Spectrostar Nano Microplate Reader (BMG Labtech, Germany). Values from blank wells were subtracted from those recorded at sample wells.

In another set of experiments, 100 µL of 2 µg/mL rhMBL (6.7 nM), CLP-1 (6.7 nM), Ficolin-1 (5 nM), Ficolin-2 (5 nM), and Ficolin-3 (3 nM), SP-A (3 nM) or SP-D (3.4 nM) in PBS were immobilized on 96-well Nunc Maxisorp Immunoplates (Costar, USA) overnight at 4°C. Plates were blocked with 200 µL of 2% BSA-TBST-Ca^2+^ for 2 h at 37°C. After washes, 100 µL of biotinylated SARS-CoV-2 S protein was added at different concentrations to the plates for 1 h at 37°C. Following washes, HRP-conjugated streptavidin (1:10000, Biospa) was incubated for 1 h at 37°C. Specific binding was detected by TMB development, as described above.

For competition-based experiments, biotinylated SARS-CoV-2 S protein was captured on 96-wells Neutravidin coated plates for 1 h at 37°C. Plates were then incubated for 1 h at 37°C with 100 µL rhMBL (0.25 µg/mL- 0.83 nM) alone or in the presence of various concentrations of D-mannose or N-acetyl-glucosamine, or D-glucose (Sigma Aldrich). Bound MBL was detected by incubation with rabbit anti-MBL antibody, followed by HRP-conjugated secondary antibody and TMB development, as described above.

For the experiments on the interaction between PTX3 domains and SARS-CoV-2 Nucleocapsid, PTX3 and its recombinant domains were immobilized on the wells of a 96-well Nunc Maxisorp plate. Then, different concentrations of biotinylated SARS-CoV-2 Nucleocapsid protein were added over the captured proteins for 1 h at 37°C. Detection of binding was achieved through incubation with HRP-conjugated streptavidin, as detailed above

### Surface plasmon resonance (SPR) studies

SPR analyses were carried out at 25 °C on a Biacore 8K instrument (GE Healthcare). MBL was immobilized on the surface of a CM5 sensor chip through standard amine coupling.

Briefly, after activation of the surface with a mixture of 1-Ethyl-3-(3-dimethylaminopropyl) carbodiimide hydrochloride and N-Hydroxysuccinimide, MBL was diluted at 50 nM in 10 mM sodium acetate buffer, pH 4.5 and injected over the surface (flow rate 10 µl/min). Free activated sites were blocked by flowing 1 M Ethanolamine, pH 8.5. Final MBL immobilization levels were around 4500 Resonance Units (RU, with 1 RU = 1 pg/mm^2^). A second surface was prepared in parallel with the same procedure, but without any ligand, and used as reference. Recombinant RBD and trimeric Spike were produced in EXPI293 cells and purified as reported (De Gasparo et al., 2021). Increasing concentration of SARS-CoV-2 RBD or Spike protein (2.5, 7.4, 22, 67, 200 and 600 nM) were injected using a single-cycle kinetics setting at a flow rate of 30 µl/min; dissociation was followed for 10 minutes. The running buffer (also used to dilute samples) was 10 mM Tris-buffered saline, pH 7.4, containing 150 mM NaCl, 2 mM CaCl_2_ and 0.005% Tween-20. In another set of experiments, the interaction was analyzed using the running buffer without CaCl_2_. Analyte responses were corrected for unspecific binding and buffer responses through the use of reference channels. Binding kinetics were determined by fitting of the experimental curves with the Langmuir 1:1 model according to standard procedures; data analyses were performed with Biacore™ Insight Evaluation Software v2.0.15.12933. In the presence of CaCl_2_, trimeric Spike bound to MBL with K_a_ (1/Ms)= 2.1e^+^4, K_d_ (1/s)= 7.3e^-^4 and K_D_=34 nM.

### Computational modeling of the MBL2 SARS-CoV-2 Spike interaction

The model of the MBL2 trimer (UniProt(Consortium, 2020) P11226) was created starting from the crystal structure of human mannose binding protein(Sheriff et al., 1994) (PDB code 1HUP). The N-terminus of MBL2 was modeled as collagen, based on the template crystal structure of collagen triple helix model(Berisio et al., 2002) (PDB code 1K6F). The binding site of mannose molecules was determined aligning the MBL2 structure to the crystal structure of rat mannose protein A(Ng et al., 2002) (PDB code 1KX1). Reference distances (∼ 40 Å) between mannose molecules were computed in PYMOL.

Putative binding sites of MBL2 were determined identifying all triplets of N-glycosylation sites at a distance between 35 Å and 50 Å in the closed state SARS-CoV-2 (Casalino et al., 2020). Distances were computed using the program ALMOST(Fu et al., 2014).

### Pseudotyped virus production

Human embryonic kidney 293T cells were transfected with a lentiviral vector expressing the Green Fluorescent Protein (GFP) under the control of a human Phosphoglycerate Kinase promoter (PGK) (Cesana et al., 2014) and a pCMV expressing vector containing the SARS-CoV-2 Spike sequence (accession number MN908947) that was codon-optimized for human expression and contained a deletion at the 3’ end aimed at deleting 19 amino acid residues at the C-terminus. An HIV *gag-pol* packaging construct and a rev-encoding plasmid were co-transfected by calcium phosphate for the production of infective viral particles. 16 h after transfection, the medium was replaced and 30 h later, supernatant was collected, filtered through 0.22 µm pore nitrocellulose filter and viral particles were pelleted by ultracentrifugation. As control, lentivirus particles were pseudotyped with the VSV-g glycoprotein that allows a high efficiency infection independently of binding to ACE2.

### Pseudotyped lentivirus binding assay

96-well Nunc Maxisorp Immunoplates (Costar, USA) were coated with 100 µL of rhMBL (3 and 1 µg/mL – 10 and 3.4 nM diluted in PBS). After overnight incubation, plates were blocked with 2% BSA diluted in TBST-Ca^2+^ for 1 h at 37°C, washed three times with TBST-Ca^2+^ and incubated for 1 h with 100 µL of SARS-CoV-2 Spike protein-pseudotyped lentivirus or VSV-pseudotyped lentivirus (ranging fom 0.1 to 1 µg/mL diluted in TBST- Ca^2+^). After washing, bound pseudotyped virus particles were lysed with 0.5% Triton X-100 and HIV p24 core protein was detected by ELISA (Perkin Elmer; USA).

### Complement deposition assay

100 µL of biotinylated SARS-CoV-2 Spike protein (either active trimer or non-covalent trimer, 1µg/mL in PBS) were captured on 96 well plates for 1 h at 37°C. After washing, wells were incubated for 1 h at 37°C with either 10% normal human serum (NHS, ComplemenTech Inc, USA), 10% C1q-depleted serum (C1qDHS), 10% C4-depleted serum (C4DHS) reconstituted or not with 25 µg/mL purified C4 (Calbiochem, USA). 10% heat-inactivated human serum (30’ at 56°C, HI-NHS) and 10% C3-depleted serum (C3DHS) were used as negative control. All the sera were diluted in 10 mM Tris-buffered saline containing 0.5 mM MgCl2, 2 mM CaCl2 and 0.05% Tween-20, also used as washing buffer. For MBL immunodepletion, 10% NHS was incubated overnight with 0.6 µg/mL rabbit anti-MBL antibody. Bound MBL-antibody complexes were separated by Dynabeads Protein G (Thermo Fisher Scientific), and the supernatant (termed MBL-ID) was used in the assay (final concentration, 10%). After washing, C5b-9 deposition was assayed by incubation for 1 h at 37°C with rabbit anti-sC5b-9 antibody (ComplemenTech Inc., Usa) diluted 1:2000 in washing buffer as described before (Stravalaci et al., 2020), followed by specific HRP-conjugated secondary antibody incubation and TMB development.

### Cell Lines

The Vero and Vero E6 cell line was obtained from the Istituto Zooprofilattico of Brescia, Italy, and ATCC, respectively. Cells were maintained in Eagle’s minimum essential medium (EMEM; Lonza) supplemented with 10% fetal bovine serum (FBS; Euroclone) and penicillin-streptomycin (complete medium).

Human embryonic kidney 293T cells, a continuous human embryonic kidney cell line containing the mutant gene of SV40 Large T Antigen (ATCC code CRL-3216), were cultured as described (Follenzi et al., 2000).

The human lung epithelial Calu-3 cell line was obtained from NovusPharma. Cells were grown in EMEM supplemented with 20% FBS and penicillin-streptomycin (complete medium).

### Human Bronchial Epithelial Cells (HBEC)

The isolation, culture, and differentiation of primary human bronchial epithelial cells (HBECs) were performed as previously reported (Scudieri et al., 2012), with some modifications. In brief, epithelial cells were obtained from mainstem human bronchi, derived from individuals undergoing lung transplant. For the present study, cells were obtained from three donors (BE37, BE63 and BE177). Epithelial cells were detached by overnight treatment of bronchi with protease XIV and then were cultured in a serum-free medium (LHC9 mixed with RPMI 1640, 1:1) containing supplements, as described (Scudieri et al., 2012). The collection of bronchial epithelial cells and their study to investigate airway epithelium physiopathology were specifically approved by the Ethics Committee of the Istituto Giannina Gaslini following the guidelines of the Italian Ministry of Health (registration number: ANTECER, 042-09/07/2018). Each patient provided informed consent to the study using a form that was also approved by the Ethics Committee.

To obtain differentiated epithelia, cells were seeded at high density (5x10^5^ cell/snapwell) on 12-mm diameter porous membranes (Snapwell inserts, Corning, code 3801). After 24 hours, the serum-free medium was removed from both sides and, on the basolateral side only, replaced with Pneumacult ALI medium (StemCell Technologies) and differentiation of cells (for 3 weeks) was performed in air-liquid interface (ALI) condition.

### Entry assay with SARS-CoV-2 Spike- pseudotyped lentivirus particles

293T cells were engineered to overexpress the SARS-CoV-2 entry receptor by transduction of a lentiviral vector expressing ACE2 (kindly provided by Massimo Pizzato, University of Trento). Lentiviral vector stock expressing ACE2 was produced as described above. The entry assay was then optimized in 96-well plate by seeding 5x10^4^ ACE2 overexpressing 293T cells/well. Twenty-four h later, cells and SARS-CoV-2 Spike-pseudotyped lentivirus stock (1:500) were incubated with serial dilutions of soluble innate immunity molecules for 30 min. The SARS-CoV-2 Spike-pseudotyped was added to the cells and forty-eight h later, cells were treated with accutase, in order to detach them from the wells, fixed and analyzed for GFP expression by cytofluorimetry.

### SARS-CoV-2 viral isolates

The SARS-CoV-2 isolate of the B.1 lineage with the Spike D614G mutation (GISAID accession ID: EPI_ISL_413489) was obtained from the nasopharyngeal swab of a mildly symptomatic patient by inoculation of Vero E6 cells as described (Clementi et al., 2020; Mycroft-West et al., 2020). The SARS-CoV-2 isolate of the South African B1.351 lineage (GISAID accession ID: EPI_ISL_1599180) was obtained from the nasopharyngeal swab of an Italian 80-year-old male patient. The SARS-CoV-2 isolate of the B1.1.7 lineage (GISAID accession ID: EPI_ISL_1924880) was obtained from the nasopharyngeal swab of an Italian 58-year-old female patient. Secondary viral stocks were generated by infection of adherent Vero E6 cells seeded in a 25 cm^2^ tissue culture flask with 0.5 ml of the primary isolate diluted in 5 ml of complete medium. Three days after infection, the supernatant was harvested and, after centrifugation, passed through a 0.45 µm filter. Aliquots of the secondary SARS-CoV-2 isolate were maintained at -80 °C. A plaque-forming assay was performed to determine viral titers.

### Infections

Calu-3 cells were seeded in 48-well plates (Corning) at the concentration of 5x10^4^ cell/well in complete medium 24 h prior to infection. Ten-fold serial dilutions of MBL (from 0.01 to 10 µg/ml – 0.034-34 nM) were incubated for 1 h with aliquots of SARS-CoV-2 containing supernatant to obtain a multiplicity of infection (MOI) of either 0.1 or 1 before incubation with Calu-3 cells (Virus+MBL). After 48 and 72 h post-infection (PI), cell culture supernatants were collected and stored at – 80 °C until determination of the viral titers by a plaque-forming assay in Vero cells.

Virus incubation with MBL was also combined with incubation of target cells. Briefly, virus incubation with MBL was performed as described above whereas Calu-3 cells were incubated with 10-fold serial dilutions of MBL (from 0.01 to 10 µg/ml – 0.034-34 nM). After 1 h, virus suspensions incubated with serial dilutions of MBL were added to MBL-treated cells (Virus+ Cells+MBL). After 48 and 72 h POST, cell culture supernatants were collected and stored at – 80 °C until determination of the viral titers by a plaque-forming assay in Vero cells. Forty-eight h before infection, the apical surface of HBEC was washed with 500 μl of PBS for 1.5 hours at 37°C, and the cultures were moved into fresh air liquid interface media. Immediately before infection, apical surfaces were washed twice to remove accumulated mucus with 500 μl of PBS with each wash lasting 30 min at 37°C. Two concentrations of either PTX3 or MBL were added to the apical surface for 1 h prior to the addition of 100 μl of viral inoculum (SARS-CoV-2) at a MOI of 1. HBEC were incubated for 2 h at 37°C. Viral inoculum was then removed and the apical surface of the cultures was washed three times with 500 μl of PBS. Cultures were incubated at 37°C for 72 h PI. Infectious virus produced by the HBEC was collected by washing the apical surface of the culture with 100 μl of PBS every 24 h up to 72 h PI. Apical washes were stored at −80°C until analysis and titered by plaque assay in Vero cells. At 72 h PI, cells were fixed in 4% paraformaldehyde for immunofluorescence analysis. All infection experiments were performed in a biosafety level 3 (BSL-3) laboratory at the Laboratory of Medical Microbiology and Virology, Vita-Salute San Raffaele University.

### Chemokine quantification

Half of the ALI medium (1 ml) was collected from each well of the lower chamber every 24 h PI and replaced with fresh ALI medium. The harvested medium was stored at -70 °C until analysis. Prior to chemokine quantification, 250 µl of medium was treated with 27 µl of Triton X-100 and heated for 30 min at 56 °C to inactivate SARS-CoV-2 infectivity.

Chemokines (IL-8 and CXCL5) were quantified by ELISA (Quantikine ELISA kits, code DY208, DY254, R&D Systems).

### Plaque-forming assay

In order to measure the virus titer of the viral stocks, a plaque-forming assay was optimized in Vero cells. Briefly, confluent Vero cells (1.5x10^6^ cell/well) seeded in 6-well plates (Corning) were incubated in duplicate with 1 ml of EMEM supplemented with 1% FBS containing 10-fold serial dilutions of SARS-CoV-2 stock. After 1 h of incubation, the viral inoculum was removed and methylcellulose (Sigma; 1 ml in EMEM supplemented with 5% FBS) was overlaid on each well. After 4 days of incubation, the cells were stained with 1% crystal violet (Sigma) in 70% methanol. The plaques were counted after examination with a stereoscopic microscope (SMZ-1500; Nikon Instruments) and the virus titer was calculated in terms of plaque forming units (PFU)/ml.

In order to determine the viral titers of the supernatant collected from Calu-3 cells at 48 and 72 h PI, confluent Vero cells (2.5x10^5^ cell/well) were seeded in 24-well plates (Corning) 24 h prior to infection. Then, cells were incubated with 300 µl of EMEM supplemented with 1% FBS containing serially diluted (1:10) virus-containing supernatants. The plaque-forming assay was performed as described above.

### Confocal and STED super-resolution microscopy

After 4% PFA fixation, HBEC cultures were incubated for 1 h with PBS and 0.1% Triton X-100 (Sigma-Aldrich), 5% normal donkey serum (Sigma-Aldrich), 2% BSA, 0.05% Tween (blocking buffer). Cells were then incubated for 2 h in blocking buffer with the following primary antibodies: mouse anti-cytokeratin 14 (Krt14) (#LL002; 1µg/ml; cat. N° 33- 168, ProSci-Incorporated, US); rabbit polyclonal anti-Spike protein (944-1218aa) (2µg/ml; cat. N° 28867-1-AP, Proteintech^®^, Germany); rat anti-MBL (#8G6; 1µg/ml; cat. N°HM1035, Hycult^®^Biotech) and rat anti-MBL (#14D12; 1µg/ml; cat. N°HM1038, Hycult^®^Biotech). After washing with PBS and 0.05% Tween, cells were incubated for 1 h with the following species- specific cross-adsorbed secondary antibodies form Invitrogen-ThermoFisher Scientific: donkey anti-rabbit IgG Alexa Fluor^®^ 488 (1µg/ml; cat. N° A-21206); donkey anti-rat IgG Alexa Fluor^®^ 594 (1µg/ml; cat. N° A-21209); donkey anti-mouse IgG Alexa Fluor^®^ 647 (1µg/ml; cat. N° A-31571). 4′,6-diamidino-2-phenylindole (DAPI) (Invitrogen) was used for nucleus staining. Cells were mounted with Mowiol^®^ (Sigma-Aldrich) and analyzed with a Leica SP8 STED3X confocal microscope system equipped with a Leica HC PLAPO CS2 63X/1.40 oil immersion lens. Confocal images (1.024 X 1.024 pixels) were acquired in *XYZ* and tiling modality (0.25 µm slice thickness) and at 1 Airy Unit (AU) of lateral resolution (pinhole aperture of 95.5µm) at a frequency of 600Hz in bidirectional mode. Alexa Fluor 488^®^ was excited with a 488 nm argon laser and emission collected from 505 to 550nm. Alexa Fluor 594^®^ was excited with a 594/604nm-tuned white light laser and emission collected from 580 to 620nm. Alexa Fluor 5647^®^ was excited with a 640/648nm-tuned white light laser and emission collected from 670 to 750nm. Frame sequential acquisition was applied to avoid fluorescence overlap. A gating between 0.4 and 7ns was applied to avoid collection of reflection and autofluorescence. 3D STED analysis was performed using the same acquisition set-up. A 660 nm CW-depletion laser (30% of power) was used for excitations of Alexa Fluor 488^®^ (Spike signal) and Alexa Fluor 594^®^ (MBL signal). STED images were acquired with a Leica HC PL APO 100×/1.40 oil STED White objective at 572.3 milli absorption unit (mAU). CW-STED and gated CW-STED were applied to Alexa Fluor 488^®^ and Alexa Fluor 594^®^, respectively. Confocal images were processed, 3D rendered and analyzed as colocalization rate between Spike and MBL with Leica Application Suite X software (LASX; version 3.5.5.19976) and presented as medium intensity projection (MIP). STED images were de-convolved with Huygens Professional software (Scientific Volume Imaging B. V.; version 19.10) and presented as MIP.

### Patient cohorts for genetic analyses

For association analyses, we investigated a total of 2,000 individuals. These included: i) 332 patients with severe COVID-19, which was defined as hospitalization with respiratory failure and a confirmed SARS-CoV-2 viral RNA PCR test from nasopharyngeal swabs. Patients were recruited from intensive care units and general wards at two hospitals in the Milan area, i.e., the Humanitas Clinical and Research Center, IRCCS, in Rozzano, Italy (140 patients); and San Gerardo Hospital, in Monza, Italy (192 patients); ii) 1,668 controls from the general Italian population with unknown COVID-19 status.

Details on DNA extraction, array genotyping and quality checks are reported elsewhere (Myocardial Infarction Genetics et al., 2009; Severe Covid et al., 2020). The dataset for cases was submitted to the European Bioinformatics Institute (www.ebi.ac.uk/gwas) under accession numbers GCST90000255 and GCST90000256, whereas the dataset for controls is deposited in the Genotypes and Phenotypes database (dbGaP; https://www.ncbi.nlm.nih.gov/gap/), under the phs000294.v1.p1 accession code.

Approvals for the project were obtained from the relevant ethics committees (Humanitas Clinical and Research Center, reference number, 316/20; the University of Milano-Bicocca School of Medicine, San Gerardo Hospital, reference number, 84/2020).

### Imputation

Genetic coverage was increased by performing single-nucleotide polymorphism (SNP) imputation on the genome build GRCh38 using the Michigan Imputation Server (https://imputation.biodatacatalyst.nhlbi.nih.gov/index.html#!) and haplotypes generated by the Trans-Omics for Precision Medicine (TOPMed) program (freeze 5) (Taliun et al., 2021), for both cases and controls. In the imputation, we used the population panel “ALL” and applied the server options to filter by an imputation of R²>0.1. In the post-imputation steps, we only retained those SNPs with R²≥0.6 and minor allele frequency (MAF) ≥1%. Next, we accurately checked cases and controls for solving within-Italian relationships and for testing the possible existence of population stratification within and across batches: to this aim, we performed principal component analysis (PCA), using a LD-pruned subset of SNPs across chromosome 10 and the Plink v.1.9 package (Chang et al., 2015). The final set of analyzed variants comprised 3,425 SNPs, distributed in the *MBL2* region (the gene +/-500 kb).

### Statistical analysis

Prism GraphPad software v. 8.0 (www.graphpad.com) was used for the statistical analyses. Comparison among groups were performed using one or two-way analysis of variance (ANOVA) and the Bonferroni’s correction. Non-linear fit of transformed data was determined by using the log (agonist) vs. response (three or four parameters).

For genetic studies, case-control allele-dose association tests were performed using the PLINK v.1.9 logistic-regression framework for dosage data. Age, sex, age*age, sex*age, and the first 10 principal components from PCA were introduced in the model as covariates. Analyses were conducted always referring to the minor allele. All P values are presented as not corrected and accompanied by odds ratio (OR) and 95% confidence interval (CI); however, in the relevant table/figure, Bonferroni-corrected thresholds for significance are indicated in the footnote/legend.

Haplotype analysis was performed in two ways: i) by selecting relevant SNPs and using the –hap-logistic option implemented in PLINK v.1.07 (Purcell et al., 2007); ii) by an unsupervised approach by means of the Beagle software v3.3 (http://faculty.washington.edu/browning/beagle/b3.html), which uses the method described by Browning & Browning (Browning and Browning, 2007) for inferring haplotype phase. In this case, we used the default setting of 1,000 permutations for calculating corrected P values.

In the meta-analysis, we took advantage of association data deposited in the Regeneron– Genetic Center database (https://rgc-covid19.regeneron.com/home) for the GHS study (Geisinger Health System; data available for 869 cases and 112,862 controls of European ancestry). Pooled Ors and Cis were calculated using the Mantel-Haenszel model (Mantel and Haenszel, 1959)

## Supplementary information

**Supplementary Table 1.** Recombinant SARS-CoV-2 proteins used in this study.

| Proteins | Host | Cat | Company |
| --- | --- | --- | --- |
| SARS-CoV-2 S1 protein, His Tag | HEK293 | S1N-C52H4 | ACROBiosystems |
| SARS-CoV-2 S2 protein, His Tag | HEK293 | S2N-C52H5 | ACROBiosystems |
| SARS-CoV-2 S protein, His Tag, active trimer | HEK293 | SPN-C52H8 | ACROBiosystems |
| SARS-CoV-2 Nucleocapsid protein, His Tag | HEK293 | NUN-C5227 | ACROBiosystems |
| SARS-CoV-2 Envelope protein. GST, His Tag | *E.coli* | ENN-C5128 | ACROBiosystems |
| Biotinylated SARS-CoV-2 S protein, His Tag, active trimer | HEK293 | SPN-C82E3 | ACROBiosystems |
| Biotinylated SARS-CoV-2 Nucleocapsid protein, His Tag | HEK293 | NUN-C82E8 | ACROBiosystems |
| SARS-CoV-2 S protein, His Tag | HEK293 | 10549-CV | R&D Systems |
| SARS-CoV-2 Nucleocapsid protein, His Tag | HEK293 | 230-30164 | RayBiotech |
| SARS-CoV-2 S protein, His Tag | HEK293 | In house | (Andreano et al., 2020) |
| SARS-CoV-2 S protein trimer, His Tag | EXPI293F cells | In house | (De Gasparo et al., 2021) |
| SARS-CoV-2 S protein, His Tag | CHO | XLGCOV-1-PPTH | ExcellGene |
| SARS-CoV-2 (2019-nCoV) Spike RBD, His Tag | HEK293 | 40592-V08H | Sino Biological |
| SARS-CoV-2 (2019-nCoV) Spike S1+S2 ECD, His Tag | Insect cells | 40589-V08B1 | Sino Biological |
| SARS-CoV-2 (2019-nCoV) Spike S1+S2 ECD (B.1.1.7), His Tag | Insect cells | 40589-V08B6 | Sino Biological |
| SARS-CoV-2 S protein (D614G), His Tag | HEK293 | SPN-C52H3 | ACROBiosystems |
| SARS-CoV-2 S protein (B.1.1.7 variant), His Tag | HEK293 | SPN-C52H6 | ACROBiosystems |
| SARS-CoV-2 S protein (B.1.351 variant), His Tag | HEK293 | SPN-C52Hc | ACROBiosystems |
| SARS-CoV-2 S protein (B.1.1.28 variant), His Tag | HEK293 | SPN-C52Hg | ACROBiosystems |

## Supplementary Figures

**Supplementary Figure 1.**
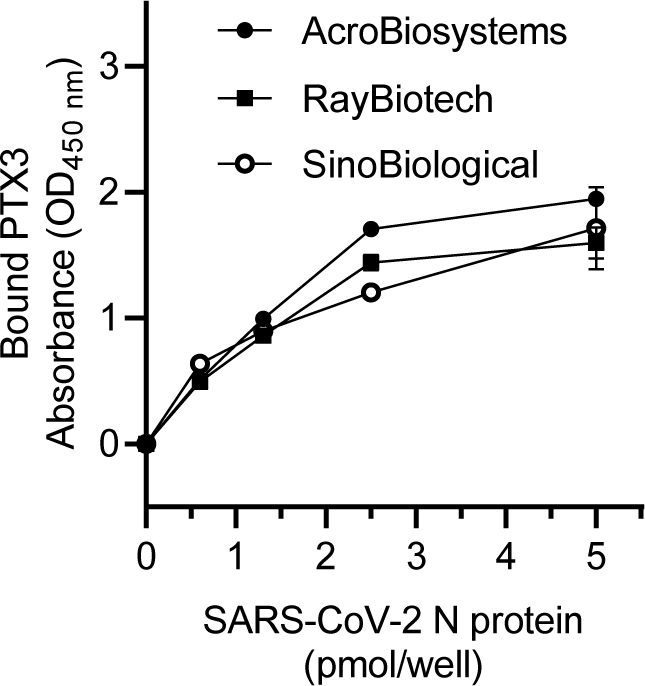
Binding of PTX3 to captured SARS-CoV-2 Nucleocapsid proteins from different companies. Data are presented as mean ± SEM of one experiment performed in duplicate.

**Supplementary Figure 2.**
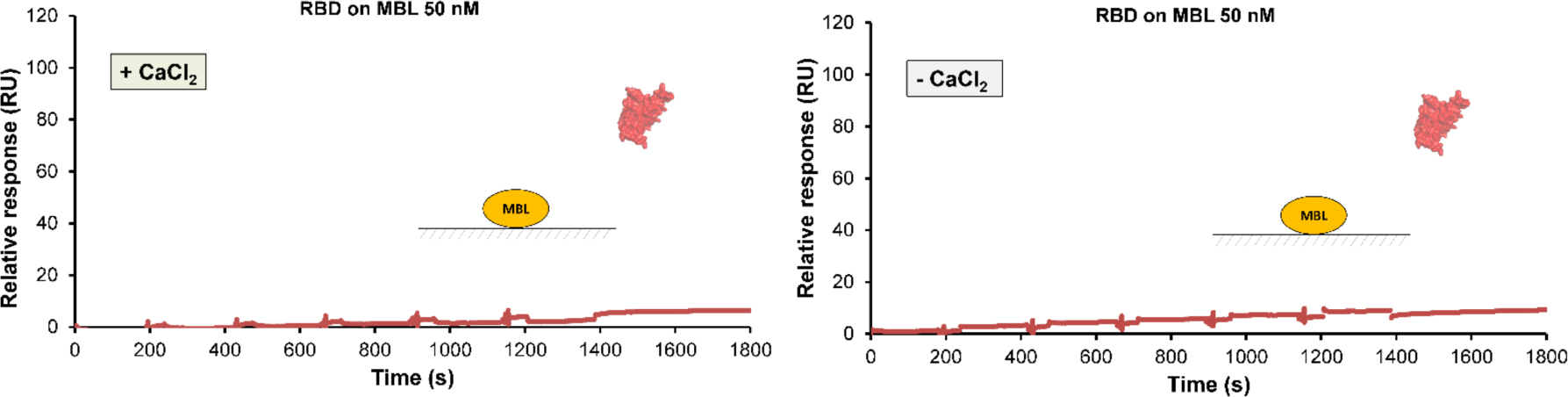
Interaction between recombinant RBD and immobilized MBL in the presence or absence of calcium, as assessed by SPR analysis.

**Supplementary Figure 3.**
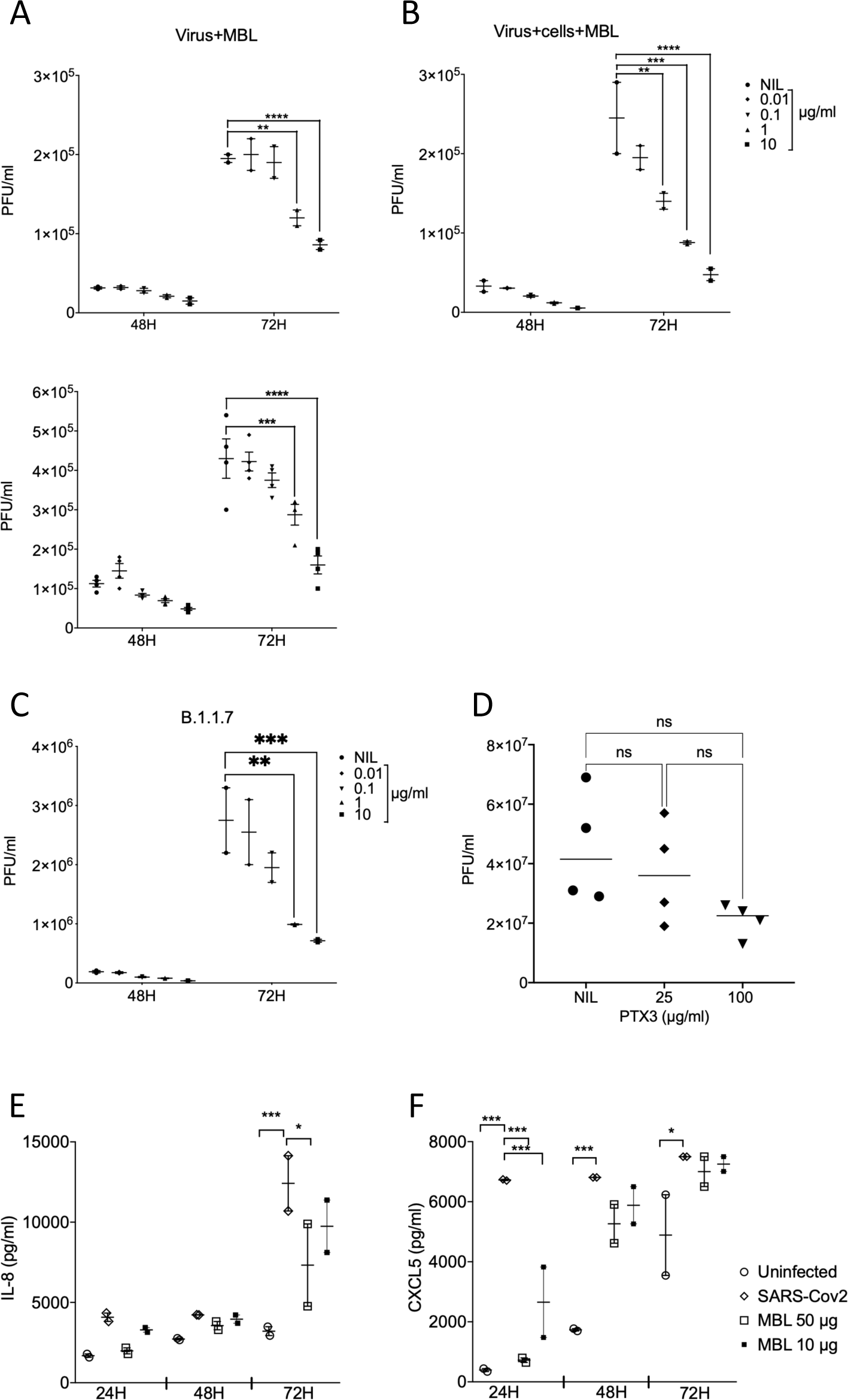
Inhibition of viral infection and chemokine production by MBL. (A-C) Inhibition of the infectivity of the D614G (isolate EPI_ISL_413489) (A, B) and B.1.1.7 SARS-CoV-2 variants by MBL in Calu-3 cells. SARS-CoV-2 (A upper panel, B and C, MOI=0.1; A lower panels, MOI=1) was preincubated in complete medium containing different concentrations of MBL (0.01–10 µg/mL– 0.034-34 nM) before incubation with Calu-3 cells (Virus + MBL) (A), or both virus and cells were pre-incubated with the same concentrations of MBL (Virus+ Cells+MBL) (B, C). After 48 and 72 h, the infectivity of SARS-CoV-2 present in cell culture supernatants was determined by a plaque-forming assay in Vero cells. NIL: no MBL. (D) SARS-CoV-2 production at the HBEC apical surface at 72 h PI, in the presence of 25 or 100 µg/ml (75 or 300 nM) PTX3. (E) Chemokine production by SARS-CoV-2 infected HBEC in the presence of MBL. Mean values of two (A, B, D) or one (C, E) experiments in duplicate cell culture are shown. ****p value <0.0001, ***p value <0.001, **p value <0.01 as determined by two-way (A, B, C, E) or one-way (D) ANOVA with Bonferroni’s correction.

**Movie S1**. **SARS-CoV-2 S protein and MBL colocalized on infected HBEC cells**. 3D rendering showing a blended reconstruction of the colocalization between SARS-CoV-2 S protein and MBL in HBEC cultures, preferentially associated to the apical side.

## References

1. Andreano, E., Piccini, G., Licastro, D., Casalino, L., Johnson, N.V., Paciello, I., Monego, S.D., Pantano, E., Manganaro, N., Manenti, A., et al. (2020). SARS-CoV-2 escape in vitro from a highly neutralizing COVID-19 convalescent plasma. bioRxiv. doi.org/10.1101/2020.12.28.424451.

2. Berisio, R., Vitagliano, L., Mazzarella, L., and Zagari, A. (2002). Crystal structure of the collagen triple helix model [(Pro-Pro-Gly)(10)](3). Protein Sci 11, 262–270.

3. Bottazzi, B., Doni, A., Garlanda, C., and Mantovani, A. (2010). An integrated view of humoral innate immunity: pentraxins as a paradigm. Annu Rev Immunol 28, 157–183.

4. Bottazzi, B., Vouret-Craviari, V., Bastone, A., De Gioia, L., Matteucci, C., Peri, G., Spreafico, F., Pausa, M., D’Ettorre, C., Gianazza, E., et al. (1997). Multimer formation and ligand recognition by the long pentraxin PTX3. Similarities and differences with the short pentraxins C-reactive protein and serum amyloid P component. J Biol Chem 272, 32817–32823.

5. Bozza, S., Bistoni, F., Gaziano, R., Pitzurra, L., Zelante, T., Bonifazi, P., Perruccio, K., Bellocchio, S., Neri, M., Iorio, A.M., et al. (2006). Pentraxin 3 protects from MCMV infection and reactivation through TLR sensing pathways leading to IRF3 activation. Blood 108, 3387–3396.

6. Browning, S.R., and Browning, B.L. (2007). Rapid and accurate haplotype phasing and missing-data inference for whole-genome association studies by use of localized haplotype clustering. Am J Hum Genet 81, 1084–1097.

7. Brunetta, E., Folci, M., Bottazzi, B., De Santis, M., Gritti, G., Protti, A., Mapelli, S.N., Bonovas, S., Piovani, D., Leone, R., et al. (2021). Macrophage expression and prognostic significance of the long pentraxin PTX3 in COVID-19. Nat Immunol 22, 19–24.

8. Carvelli, J., Demaria, O., Vely, F., Batista, L., Chouaki Benmansour, N., Fares, J., Carpentier, S., Thibult, M.L., Morel, A., Remark, R., et al. (2020). Association of COVID-19 inflammation with activation of the C5a-C5aR1 axis. Nature 588, 146–150.

9. Casalino, L., Gaieb, Z., Goldsmith, J.A., Hjorth, C.K., Dommer, A.C., Harbison, A.M., Fogarty, C.A., Barros, E.P., Taylor, B.C., McLellan, J.S., et al. (2020). Beyond Shielding: The Roles of Glycans in the SARS-CoV-2 Spike Protein. ACS Cent Sci 6, 1722–1734.

10. Cesana, D., Ranzani, M., Volpin, M., Bartholomae, C., Duros, C., Artus, A., Merella, S., Benedicenti, F., Sergi Sergi, L., Sanvito, F., et al. (2014). Uncovering and dissecting the genotoxicity of self-inactivating lentiviral vectors in vivo. Mol Ther 22, 774–785.

11. Chang, C.C., Chow, C.C., Tellier, L.C., Vattikuti, S., Purcell, S.M., and Lee, J.J. (2015). Second-generation PLINK: rising to the challenge of larger and richer datasets. Gigascience 4, 7.

12. Chiodo, F., Bruijns, S.C.M., Rodriguez, E., Li, R.J.E., Molinaro, A., Silipo, A., Di Lorenzo, F., Garcia-Rivera, D., Valdes-Balbin, Y., Verez-Bencomo, V., et al. (2020). Novel ACE2-Independent Carbohydrate-Binding of SARS-CoV-2 Spike Protein to Host Lectins and Lung Microbiota. bioRxiv, doi.org/10.1101/2020.05.13.092478.

13. Chu, H., Chan, J.F., Yuen, T.T., Shuai, H., Yuan, S., Wang, Y., Hu, B., Yip, C.C., Tsang, J.O., Huang, X., et al. (2020). Comparative tropism, replication kinetics, and cell damage profiling of SARS-CoV-2 and SARS-CoV with implications for clinical manifestations, transmissibility, and laboratory studies of COVID-19: an observational study. Lancet Microbe 1, e14–e23.

14. Clementi, N., Criscuolo, E., Diotti, R.A., Ferrarese, R., Castelli, M., Dagna, L., Burioni, R., Clementi, M., and Mancini, N. (2020). Combined Prophylactic and Therapeutic Use Maximizes Hydroxychloroquine Anti-SARS-CoV-2 Effects in vitro. Front Microbiol 11, 1704.

15. Consortium, T.U. (2020). UniProt: the universal protein knowledgebase in 2021. Nucleic Acids Res 49, D480–D489.

16. De Gasparo, R., Pedotti, M., Simonelli, L., Nickl, P., Muecksch, F., Cassaniti, I., Percivalle, E., Lorenzi, J.C.C., Mazzola, F., Magri, D., et al. (2021). Bispecific IgG neutralizes SARS-CoV-2 variants and prevents escape in mice. Nature 593, 424–428.

17. Desmyter, J., Melnick, J.L., and Rawls, W.E. (1968). Defectiveness of interferon production and of rubella virus interference in a line of African green monkey kidney cells (Vero). J Virol 2, 955–961.

18. Fajgenbaum, D.C., and June, C.H. (2020). Cytokine Storm. N Engl J Med 383, 2255–2273.

19. Follenzi, A., Ailles, L.E., Bakovic, S., Geuna, M., and Naldini, L. (2000). Gene transfer by lentiviral vectors is limited by nuclear translocation and rescued by HIV-1 pol sequences. Nat Genet 25, 217–222.

20. Fu, B., Sahakyan, A.B., Camilloni, C., Tartaglia, G.G., Paci, E., Caflisch, A., Vendruscolo, M., and Cavalli, A. (2014). ALMOST: an all atom molecular simulation toolkit for protein structure determination. J Comput Chem 35, 1101–1105.

21. Gao, T., Hu, M., Zhang, X., Li, H., Zhu, L., Liu, H., Dong, Q., Zhang, Z., Wang, Z., Hu, Y., et al. (2020). Highly pathogenic coronavirus N protein aggravates lung injury by MASP-2-mediated complement over-activation. medRxiv, 10.1101/2020.1103.1129.20041962.

22. Garlanda, C., Bottazzi, B., Magrini, E., Inforzato, A., and Mantovani, A. (2018). PTX3, a Humoral Pattern Recognition Molecule, in Innate Immunity, Tissue Repair, and Cancer. Physiol Rev 98, 623–639.

23. Garred, P., Pressler, T., Lanng, S., Madsen, H.O., Moser, C., Laursen, I., Balstrup, F., Koch, C., and Koch, C. (2002). Mannose-binding lectin (MBL) therapy in an MBL-deficient patient with severe cystic fibrosis lung disease. Pediatr Pulmonol 33, 201–207.

24. Han, B., Ma, X., Zhang, J., Zhang, Y., Bai, X., Hwang, D.M., Keshavjee, S., Levy, G.A., McGilvray, I., and Liu, M. (2012). Protective effects of long pentraxin PTX3 on lung injury in a severe acute respiratory syndrome model in mice. Lab Invest 92, 1285–1296.

25. Hartenian, E., Nandakumar, D., Lari, A., Ly, M., Tucker, J.M., and Glaunsinger, B.A. (2020). The molecular virology of coronaviruses. J Biol Chem 295, 12910–12934.

26. Holmskov, U., Thiel, S., and Jensenius, J.C. (2003). Collections and ficolins: humoral lectins of the innate immune defense. Annu Rev Immunol 21, 547–578.

27. Ip, W.K., Chan, K.H., Law, H.K., Tso, G.H., Kong, E.K., Wong, W.H., To, Y.F., Yung, R.W., Chow, E.Y., Au, K.L., et al. (2005). Mannose-binding lectin in severe acute respiratory syndrome coronavirus infection. J Infect Dis 191, 1697–1704.

28. Jensenius, J.C., Jensen, P.H., McGuire, K., Larsen, J.L., and Thiel, S. (2003). Recombinant mannan-binding lectin (MBL) for therapy. Biochem Soc Trans 31, 763–767.

29. Karwaciak, I., Salkowska, A., Karas, K., Dastych, J., and Ratajewski, M. (2021). Nucleocapsid and Spike Proteins of the Coronavirus SARS-CoV-2 Induce IL6 in Monocytes and Macrophages-Potential Implications for Cytokine Storm Syndrome. Vaccines (Basel) 9, 54.

30. King, C., and Sprent, J. (2021). Dual Nature of Type I Interferons in SARS-CoV-2- Induced Inflammation. Trends Immunol 42, 312–322.

31. Koch, A., Melbye, M., Sorensen, P., Homoe, P., Madsen, H.O., Molbak, K., Hansen, C.H., Andersen, L.H., Hahn, G.W., and Garred, P. (2001). Acute respiratory tract infections and mannose-binding lectin insufficiency during early childhood. JAMA 285, 1316–1321.

32. Lempp, F.A., Soriaga, L., Montiel-Ruiz, M., Benigni, F., Noack, J., Park, Y.-J., Bianchi, S., Walls, A.C., Bowen, J.E., Zhou, J., et al. (2021). Membrane lectins enhance SARS-CoV-2 infection and influence the neutralizing activity of different classes of antibodies. bioRxiv, doi.org/10.1101/2021.04.03.438258.

33. Lipscombe, R.J., Sumiya, M., Hill, A.V., Lau, Y.L., Levinsky, R.J., Summerfield, J.A., and Turner, M.W. (1992). High frequencies in African and non-African populations of independent mutations in the mannose binding protein gene. Hum Mol Genet 1, 709–715.

34. Lu, Q., Liu, J., Zhao, S., Gomez Castro, M.F., Laurent-Rolle, M., Dong, J., Ran, X., Damani-Yokota, P., Tang, H., Karakousi, T., et al. (2021). SARS-CoV-2 exacerbates proinflammatory responses in myeloid cells through C-type lectin receptors and Tweety family member 2. Immunity. doi: 10.1016/j.immuni.2021.05.006. In press.

35. Madsen, H.O., Garred, P., Kurtzhals, J.A., Lamm, L.U., Ryder, L.P., Thiel, S., and Svejgaard, A. (1994). A new frequent allele is the missing link in the structural polymorphism of the human mannan-binding protein. Immunogenetics 40, 37–44.

36. Madsen, H.O., Garred, P., Thiel, S., Kurtzhals, J.A., Lamm, L.U., Ryder, L.P., and Svejgaard, A. (1995). Interplay between promoter and structural gene variants control basal serum level of mannan-binding protein. J Immunol 155, 3013–3020.

37. Mantel, N., and Haenszel, W. (1959). Statistical aspects of the analysis of data from retrospective studies of disease. J Natl Cancer Inst 22, 719–748.

38. McBride, R., van Zyl, M., and Fielding, B.C. (2014). The coronavirus nucleocapsid is a multifunctional protein. Viruses 6, 2991–3018.

39. Medetalibeyoglu, A., Bahat, G., Senkal, N., Kose, M., Avci, K., Sayin, G.Y., Isoglu- Alkac, U., Tukek, T., and Pehlivan, S. (2021). Mannose binding lectin gene 2 (rs1800450) missense variant may contribute to development and severity of COVID-19 infection. Infect Genet Evol 89, 104717.

40. Merad, M., and Martin, J.C. (2020). Pathological inflammation in patients with COVID-19: a key role for monocytes and macrophages. Nat Rev Immunol 20, 355–362.

41. Mycroft-West, C.J., Su, D., Pagani, I., Rudd, T.R., Elli, S., Gandhi, N.S., Guimond, S.E., Miller, G.J., Meneghetti, M.C.Z., Nader, H.B., et al. (2020). Heparin Inhibits Cellular Invasion by SARS-CoV-2: Structural Dependence of the Interaction of the Spike S1 Receptor-Binding Domain with Heparin. Thromb Haemost 120, 1700–1715.

42. Myocardial Infarction Genetics, C., Kathiresan, S., Voight, B.F., Purcell, S., Musunuru, K., Ardissino, D., Mannucci, P.M., Anand, S., Engert, J.C., Samani, N.J., et al. (2009). Genome-wide association of early-onset myocardial infarction with single nucleotide polymorphisms and copy number variants. Nat Genet 41, 334–341.

43. Ng, K.K., Kolatkar, A.R., Park-Snyder, S., Feinberg, H., Clark, D.A., Drickamer, K., and Weis, W.I. (2002). Orientation of bound ligands in mannose-binding proteins. Implications for multivalent ligand recognition. J Biol Chem 277, 16088–16095.

44. Pairo-Castineira, E., Clohisey, S., Klaric, L., Bretherick, A.D., Rawlik, K., Pasko, D., Walker, S., Parkinson, N., Fourman, M.H., Russell, C.D., et al. (2021). Genetic mechanisms of critical illness in COVID-19. Nature 591, 92–98.

45. Purcell, S., Neale, B., Todd-Brown, K., Thomas, L., Ferreira, M.A., Bender, D., Maller, J., Sklar, P., de Bakker, P.I., Daly, M.J., et al. (2007). PLINK: a tool set for whole-genome association and population-based linkage analyses. Am J Hum Genet 81, 559–575.

46. Reading, P.C., Bozza, S., Gilbertson, B., Tate, M., Moretti, S., Job, E.R., Crouch, E.C., Brooks, A.G., Brown, L.E., Bottazzi, B., et al. (2008). Antiviral Activity of the Long Chain Pentraxin PTX3 against Influenza Viruses. J Immunol 180, 3391–3398.

47. Risitano, A.M., Mastellos, D.C., Huber-Lang, M., Yancopoulou, D., Garlanda, C., Ciceri, F., and Lambris, J.D. (2020). Complement as a target in COVID-19? Nat Rev Immunol 20, 343–344.

48. Schirinzi, A., Pesce, F., Laterza, R., D’Alise, M.G., Lovero, R., Fontana, A., Contino, R., and Di Serio, F. (2021). Pentraxin 3: Potential prognostic role in SARS-CoV-2 patients admitted to the emergency department. J Infect 82, 84–123.

49. Scudieri, P., Caci, E., Bruno, S., Ferrera, L., Schiavon, M., Sondo, E., Tomati, V., Gianotti, A., Zegarra-Moran, O., Pedemonte, N., et al. (2012). Association of TMEM16A chloride channel overexpression with airway goblet cell metaplasia. J Physiol 590, 6141–6155.

50. Severe Covid, G.G., Ellinghaus, D., Degenhardt, F., Bujanda, L., Buti, M., Albillos, A., Invernizzi, P., Fernandez, J., Prati, D., Baselli, G., et al. (2020). Genomewide Association Study of Severe Covid-19 with Respiratory Failure. N Engl J Med 383, 1522–1534.

51. Sheriff, S., Chang, C.Y., and Ezekowitz, R.A.B. (1994). Human mannose-binding protein carbohydrate recognition domain trimerizes through a triple α-helical coiled-coil. Nat Struct Biol 1, 789–794.

52. Stravalaci, M., Davi, F., Parente, R., Gobbi, M., Bottazzi, B., Mantovani, A., Day, A.J., Clark, S.J., Romano, M.R., and Inforzato, A. (2020). Control of Complement Activation by the Long Pentraxin PTX3: Implications in Age-Related Macular Degeneration. Front Pharmacol 11, 591908.

53. Sumiya, M., Super, M., Tabona, P., Levinsky, R.J., Arai, T., Turner, M.W., and Summerfield, J.A. (1991). Molecular basis of opsonic defect in immunodeficient children. Lancet 337, 1569–1570.

54. Taliun, D., Harris, D.N., Kessler, M.D., Carlson, J., Szpiech, Z.A., Torres, R., Taliun, S.A.G., Corvelo, A., Gogarten, S.M., Kang, H.M., et al. (2021). Sequencing of 53,831 diverse genomes from the NHLBI TOPMed Program. Nature 590, 290–299.

55. Wang, J., Jiang, M., Chen, X., and Montaner, L.J. (2020). Cytokine storm and leukocyte changes in mild versus severe SARS-CoV-2 infection: Review of 3939 COVID-19 patients in China and emerging pathogenesis and therapy concepts. J Leukoc Biol 108, 17–41.

56. Watanabe, Y., Allen, J.D., Wrapp, D., McLellan, J.S., and Crispin, M. (2020). Site-specific glycan analysis of the SARS-CoV-2 spike. Science 369, 330–333.

57. Wu, F., Zhao, S., Yu, B., Chen, Y.M., Wang, W., Song, Z.G., Hu, Y., Tao, Z.W., Tian, J.H., Pei, Y.Y., et al. (2020). A new coronavirus associated with human respiratory disease in China. Nature 579, 265–269.

58. Yuan, F.F., Tanner, J., Chan, P.K., Biffin, S., Dyer, W.B., Geczy, A.F., Tang, J.W., Hui, D.S., Sung, J.J., and Sullivan, J.S. (2005). Influence of FcgammaRIIA and MBL polymorphisms on severe acute respiratory syndrome. Tissue Antigens 66, 291–296.

59. Zeng, W., Liu, G., Ma, H., Zhao, D., Yang, Y., Liu, M., Mohammed, A., Zhao, C., Yang, Y., Xie, J., et al. (2020). Biochemical characterization of SARS-CoV-2 nucleocapsid protein. Biochem Biophys Res Commun 527, 618–623.

60. Zhang, H., Zhou, G., Zhi, L., Yang, H., Zhai, Y., Dong, X., Zhang, X., Gao, X., Zhu, Y., and He, F. (2005). Association between mannose-binding lectin gene polymorphisms and susceptibility to severe acute respiratory syndrome coronavirus infection. J Infect Dis 192, 1355–1361.

61. Zhang, Q., Bastard, P., Liu, Z., Le Pen, J., Moncada-Velez, M., Chen, J., Ogishi, M., Sabli, I.K.D., Hodeib, S., Korol, C., et al. (2020). Inborn errors of type I IFN immunity in patients with life-threatening COVID-19. Science 370, eabd4570.

62. Zhou, Y., Lu, K., Pfefferle, S., Bertram, S., Glowacka, I., Drosten, C., Pohlmann, S., and Simmons, G. (2010). A single asparagine-linked glycosylation site of the severe acute respiratory syndrome coronavirus spike glycoprotein facilitates inhibition by mannose-binding lectin through multiple mechanisms. J Virol 84, 8753–8764.

63. Zhu, N., Zhang, D., Wang, W., Li, X., Yang, B., Song, J., Zhao, X., Huang, B., Shi, W., Lu, R., et al. (2020). A Novel Coronavirus from Patients with Pneumonia in China, 2019. N Engl J Med 382, 727–733.

